# Organomics: A concept reflecting the importance of PET/CT healthy organ radiomics in non-small cell lung cancer prognosis prediction using machine learning

**DOI:** 10.1101/2024.05.15.24307393

**Authors:** Yazdan Salimi, Ghasem Hajianfar, Zahra Mansouri, Amirhosein Sanaat, Mehdi Amini, Isaac Shiri, Habib Zaidi

## Abstract

**Purpose:** Non-small cell lung cancer (NSCLC) is the most common subtype of lung cancer. Patient survival prediction using machine learning and radiomics analysis proved to provide promising outcomes. However, most studies reported in the literature focused on information extracted from malignant lesions. This study aims to explore the relevance and additional value of information extracted from healthy organs in addition to tumoral tissue using machine learning algorithms.

**Methods:** This study included PET/CT images of 154 patients collected from available online databases. The gross tumour volume (GTV) and 33 volumes of interest defined on healthy organs were segmented using nnU-Net deep learning-based segmentation. Subsequently, 107 radiomic features were extracted from PET and CT images (Organomics). Clinical information was combined with PET and CT radiomics from organs and GTVs considering 19 different combinations of inputs. Finally, different feature selection (FS, 5 methods) and machine learning (ML, 6 algorithms) algorithms were tested in a three-fold data split cross-validation scheme. The performance of the models was quantified in terms of the concordance index (C-index) metric.

**Results:** For an input combination of all radiomics information, most of the selected features belonged to PET Organomics and CT Organomics. The highest C-Index (0.68) was achieved using univariate C-Index FS method and random survival forest ML model using CT Organomics + PET Organomics as input as well as minimum depth FS method and CoxPH ML model using PET Organomics as input. Considering all 17 combinations with C-Index higher than 0.65, Organomics from PET or CT images were used as input in 16 of them.

**Conclusion:** The selected features and C-Indices demonstrated that the additional information extracted from healthy organs of both PET and CT imaging modalities improved the machine learning performance. Organomics could be a step toward exploiting the whole information available from multimodality medical images, contributing to the emerging field of digital twins in healthcare.

## Introduction

Lung cancer is the second most common cancer in all genders, while the most common subtype of lung cancer is non-small cell lung cancer (NSCLSC), a leading cause of death among other malignancies [1]. Knowledge of prognosis prior to treatment and during the treatment can be useful to change or optimize the treatment strategy or prevent other post-treatment. Radiomics information coupled with machine learning algorithms showed potential to predict the prognosis for NSCLC patients after treatment [2; 3], while most of the available studies using artificial intelligence (AI) [3-5] focused on radiomic features extracted from the tumoral region and used clinical information, such as age, gender, and blood tests as additional information. Amini et al. [5] developed machine learning models to predict survival using different image fusion strategies and radiomics extracted from the GTV on the same population [4]. Lee et al. [6] extracted peritumoral image features and reported gain in classification performance which depends on tumour size. Hosny et al. [7] showed that deep learning classification algorithms emphasized the importance of peritumoral tissue in patient risk estimation. Mattonen et al. [8] reported the importance of metabolic tumor volume penumbra extended by 1 cm in NSCLC recurrence. Guo et al. [9] evaluated the predictive value of dosiomics and CT radiomics of esophageal tumour GTV and whole oesophagus for predicting complications after radiotherapy. They reported combination of GTV and whole oesophagus as the best predictor using machine learning models. Lam et al. [10] used multi-omics data including radiomics and dosiomics extracted from eight volumes of interest irradiated around the nasopharyngeal GTV to predict the adaptive radiotherapy eligibility in nasopharyngeal cancer patients. They reported the best performance for radiomics plus dosiomics extracted from these eight regions plus the GTV. They did not compare the GTV only versus added value of the surrounding organs. Kibrom et al. [11] reported that the position of lesions relative to spleen has additional predictive value in lymphoma patients treated with radiopharmaceutical therapy. During the Covid-19 pandemic, few studies reported the importance of gastrointestinal finding in predicting patient prognosis [12; 13]. Szabo et al. [14] reported the importance of pericardiac fat in the prognostic prediction of patients with heart failure.

We believe that overall patients’ health condition may play a role in prognosis. Besides we hypothesize that it may contain some information reflecting overall patients’ health in the radiomic features space from structural (CT) and metabolic (PET) images acquired from these regions. Deep learning-based segmentation enables fast and reliable delimitation of healthy organs and hence evaluation any organ separately [15; 16]. To the best of our knowledge, the contribution of healthy organs is always overlooked and studies exploring the importance of healthy organs to estimate overall patient characteristics in survival prediction in NSCLC patients are lacking.

The aim of this study was to use as much as possible image information available from PET/CT images to predict the prognosis in terms of overall survival prediction in patients with NSCLC malignancies. We used radiomic features extracted from 33 organs and tumoral tissues and evaluated the added value of healthy organs radiomics in a comprehensive study using multiple feature selection and machine learning models. The primary question addressed was whether the incorporation of total body organ information could enhance the accuracy of AI-based predictions of overall survival.

## Materials and Methods

### Dataset

This study used the Radiogenomics NSCLC dataset downloaded from the TCIA public database [17]. Cases where PET/CT imaging data are available were separated and the DICOM images converted to NIFTI format. From 211 cases, there were 166 cases with PET/CT and after preprocessing and excluding images with any kind of processing error or missing data, a total number of 154 PET/CT images was included for training/testing. A detailed description of the demographics, acquisition and reconstruction parameters is summarized in Table 1. We calculated the time difference between the PET acquisition date and the date of the last follow up recorded on the dataset description. It should be mentioned that the PET/CT acquisition date was not available for a few cases in the metadata provided by TCIA. For these cases, the DICOM acquisition date information was used. PET images were converted to standard izeduptake value (SUV) prior to feature extraction.

**Table 1.** Demographic description of the dataset included in this study summarizing patient information, PET and CT acquisition/reconstruction parameters.

|  |  |  |
| --- | --- | --- |
| **Demographics** | Age (years) | $67.2 \pm 11.29$ |
| | Height (meters) | $1.69 \pm 0.17$ |
| | Weight (Kg) | $76.26 \pm 18.51$ |
|  | Gender | Male (# 97), Female (#57) |
|  | Affiliation | Stanford (#87), VA (# 67) |
|  | Survival status | Alive (#110), Deceased (#44) |
| **PET** | Manufacturer | Siemens (#10), GE (#144) |
| | PET spacing (mm) | $4.37 \pm 0.84$ |
| | PET Injected Activity (MBq) | $453.16 \pm 90.46$ |
| | Time per bed (minutes) | $2.33 \pm 0.85$ |
|  | Scatter correction method | Model-Based, Convolution Subtraction |
|  | PET Reconstruction Method | OSEM, 3D IR, VPFX, OSEM PSF, VPHDS |
| **CT** | kVp | 80, 100, 120, 130, 140 |
| | Pitch Factor | $1.08 \pm 0.29$ |
| | Average Tube Current (mA) | $267.58 \pm 163.93$ |

### Organs segmentation

We used extended and upgraded versions of previously trained deep learning-based segmentation models in our department [15] to segment 28 volumes of interest in healthy organs on the CT images. Those models were trained using nnU-Net [18] segmentation pipeline using five-fold data split and ensembling all five-folds inferred on the RadioGenomics CT compartment of PET/CT dataset. The 3Dfullress training model weas continued using 2000 epochs and initial learning rate of 3e-5 decreased after each epoch. The segmented organs were visually checked searching for potential outliers presenting with significant errors. The list of segmented organs is provided in Table 2.

**Table 2.** List of segmented organs for three subgroups of soft, lung, and bony tissues. LLL: left lower lobe, RLL: right lower lobe, RML: right middle lobe, LUL: left upper lobe, RUL: right upper lobe.

|  |  |  |
| --- | --- | --- |
| **Boney Structures** | 1 | Clavicles |
|  | 2 | Hips |
|  | 3 | Sacrum |
|  | 4 | Ribs |
|  | 5 | Vertebrae |
|  | 6 | Femoral Heads |
| **Soft Tissue** | 7 | Adrenal Glands |
|  | 8 | Aorta |
|  | 9 | Brain |
|  | 10 | Colon |
|  | 11 | Esophagus |
|  | 12 | Eyeballs |
|  | 13 | Whole Cardiac |
|  | 14 | Cardiac Right Atrium |
|  | 15 | Cardiac Left Atrium |
|  | 16 | Cardiac Left Ventricle Cavity |
|  | 17 | Cardiac Right Ventricle |
|  | 18 | Cardiac Left Myocardium |
|  | 19 | Kidneys |
|  | 20 | Liver |
|  | 21 | Pancreas |
|  | 22 | Rectum |
|  | 23 | Rectus Lumborum Muscles |
|  | 24 | Small Intestine |
|  | 25 | Spleen |
|  | 26 | Stomach |
|  | 27 | Urinary Bladder |
| **Lung Tissue** | 28 | Whole Lungs |
|  | 29 | Lung LLL |
|  | 30 | Lung RLL |
|  | 31 | Lung RML |
|  | 32 | Lung LUL |
|  | 33 | Lung RUL |

### GTV segmentation

We used nnU-Net pipeline to train a 3Dfullres deep learning model to segment GTV on CT of PET/CT images. We used three online available datasets including LIDC [19] (dataset #1) and NSCLC (dataset #2) and manual segmentations available on Radiogenomics [17] (same patients as PET/CT images, dataset #3) datasets for model training using a five-fold data split. The Radiogenomics dataset had the same patients whom PET/CT images were used to train the survival ML models. It should be mentioned that the Radiogenomics diagnostic CTs with available manual segmentation (143 pair of CT and GTV segmentations) were used both as part of training set and testing set. We used datasets #1 and #2 to increase the number of training datasets and gain a robust model capable of segmenting CT of PET/CT images with a lower image quality.

These three datasets were visually assessed and cases with presenting with errors were excluded from training. After exclusion, 384 cases from NSCLSC dataset, 143 cases from Radiogenomics dataset, and 787 cases from LIDC dataset (total of 1314 pairs of CT and GTV segmentation) were included. Similar to organ segmentation part, we ensembled the output from all five folds inferenced on CT images of PET/CT. The GTV segmentations were visually checked and compared with the available ground truth data provided on the diagnostic CT which was not co-registered with the PET/CT images in few cases.

### Feature extraction

We used Pyradiomics (version 3.1.0) [20] library to extract 107 radiomic features, including First-order Statistics (19 features), Shape-based (3D) (16 features), Shape-based (2D) (10 features), Gray Level Co-occurrence Matrix (24 features), Gray Level Run Length Matrix (16 features), Gray Level Size Zone Matrix (16 features), Neighboring Gray Tone Difference Matrix (5 features), and Gray Level Dependence Matrix (14 features). We clipped the images prior to feature extraction depending on organ composition for organs and used a predefined clipping value for malignant lesions. We manually classified organs in one of three subgroups, namely lung, soft-tissue, and bony structures. Then, for each category, prior to extracting the radiomic features, the images were clipped between empirical minimum and maximum values to emphasis the image histogram on the heterogeneities inside these tissues. The clipping values were -900 to 0, -300 to 300, and 0 to 800 HU for lung, soft-tissue and bony structures, respectively. PET images were clipped between 0 and 40 SUV before feature extraction for all segmentation masks.

We extracted features using bin width equal to 10 HUs and 0.4 SUV for CT and PET images, respectively. PET and CT images were resampled to 4 × 4 × 4 mm^3^ and 1.5 × 1.5 × 1.5 mm^3^, respectively, prior to feature extraction.

### Feature selection and machine learning

We considered 19 possible combinations of five input data including PET Organomics, CT Organomics, PET GTV, CT GTV, and clinical information. Table 3 summarizes these 19 strategies.

**Table 3.**
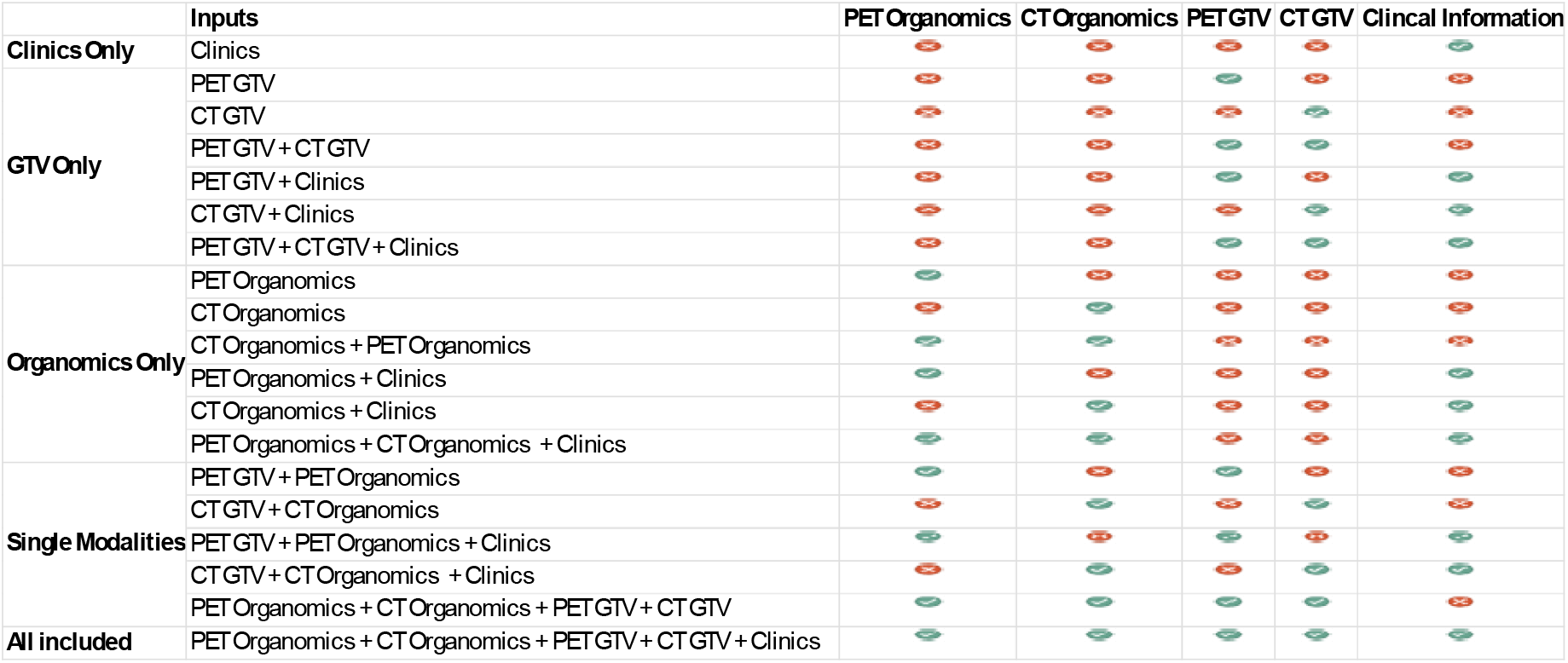
Summary of all 19 combinations of input data used in this study. Red cross sign means not used whereas the green thick sign means using that input. For better readability, they were classified in four subgroups and all included means using all five inputs as predictors.

Figure 1 shows the flowchart of steps followed in this study protocol.

**Figure 1.**
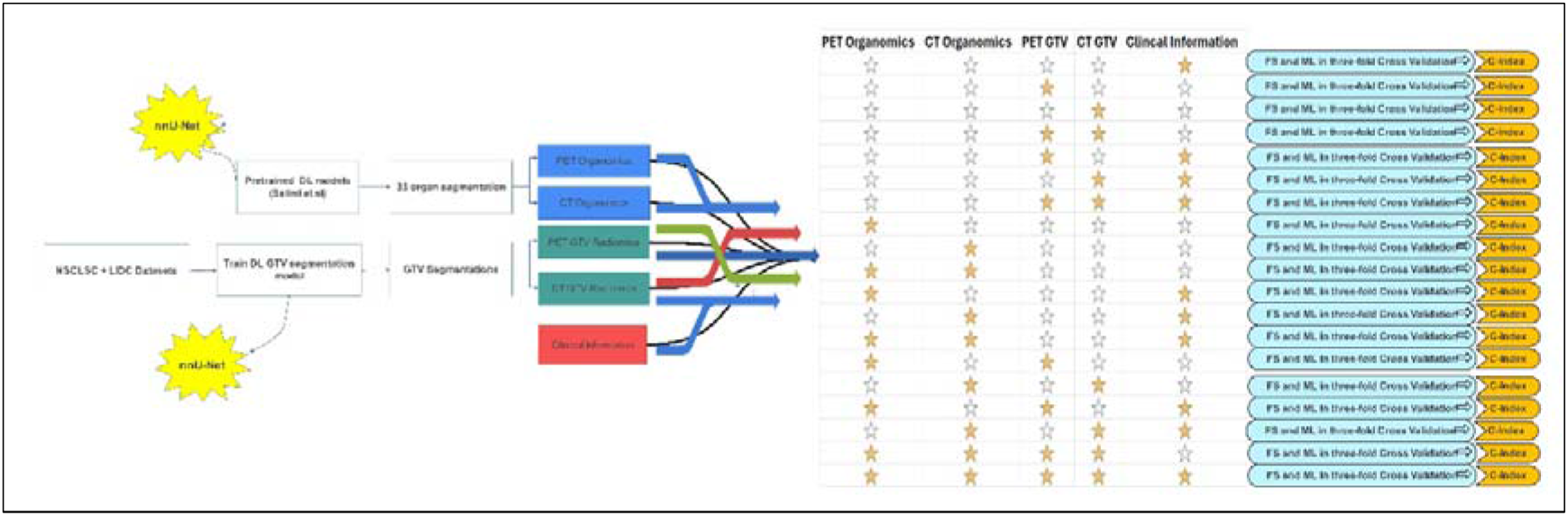
Flowchart summarizing the different steps involved in the study protocol. All 19 input combinations were trained using three-fold cross-validation data split. Filled yellow star means using that input whereas blank (white) star means that input was not used.

We used combinations of five feature selection (FS), six machine learning (ML) models and 19 types of input in three-fold data split to train overall 570 × 3 (1710) models and compared the performance in terms of concordance index (C-index). Different FS algorithms were used in this study, including Minimal Depth (MD), Mutual Information (MI), Univariate C-Index (UCI), Boruta, Variable hunting (VH), Variable hunting Variable Importance (VH.VIMP). We implemented 6 ML models, including Cox Boost (CB), Cox Proportional Hazard regression (CoxPH), Generalized Linear Model Network (GLMN), GLM Boosting (GLMB), Random Survival Forest (RSF), and Survival Tree (ST). Details about the implemented methods are provided in supplementary material.

First, we applied 3-fold nested cross-validation for each input. In each fold (external fold), we used z-score method to normalize feature values based on train dataset and transformed the values (mean and standard deviation) to test dataset. For remove redundant feature, we used Spearman correlation test with a threshold of 90%. This method removes one of the features that has a Spearman correlation coefficient over 90%. Then, FS algorithms were applied on the train dataset. The best selected features for each FS method were fed to ML algorithms. Internal 3-fold cross-validation with grid search was used for hyperparameter optimization. The detail of these parameters is provided in supplementary material. The trained model with best hyperparameter was evaluated on test dataset with 1000 bootstraps. Model evaluation was performed with C-index. Mean and standard deviation of 3000 C-indices was reported for each model. The mlr package version 2.18 in R 4.1.2 was used for model development.

### Statistical Analysis

The top performance models with respect to the C-index were selected for Kaplan-Meir (KM) curve analysis. The risk score in the test dataset for each fold KM was extracted and combined for all patients. The risk scores were transformed to high-risk and low-risk groups using the median value as the threshold. The log-rank test was used to show significant differences between two groups (p-value<0.05).

## Results

### Segmentation accuracy

Figure 2 shows an example of GTVs segmented on both diagnostic quality CT and CT of a PET/CT image for a case with Dice coefficient equal to 0.87, which is lower than the average value. An average Dice coefficient of 0.92 ± 0.08 was calculated on the 143 diagnostic cases showing excellent segmentation performance on GTV segmentation. Figure 3 presents an example of organs segmented on CT of a PET/CT image showing excellent performance of organ segmentation model as reported in a previous study [15].

**Figure 2.**
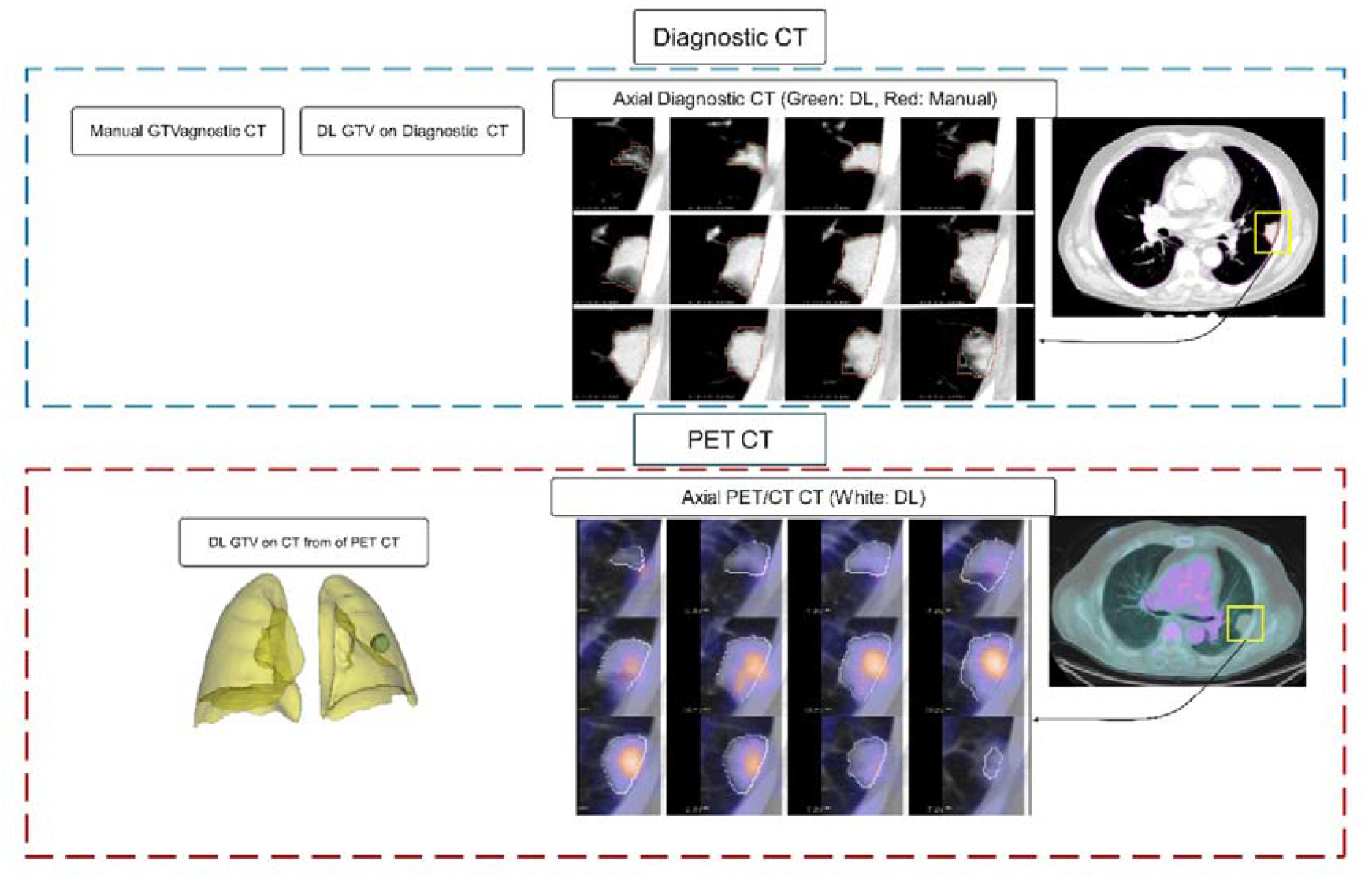
GTV segmentations for a case with Dice coefficient of 0.87 on diagnostic CT images. The top row shows a pair of manual (ground truth) and deep learning (DL) segmentation output on a diagnostic CT image where the axial magnified slices compare the manual (red) and DL (green) contours. The bottom row shows the corresponding axial slices segmented using DL on CT of a PET/CT image. The 3D visualization shows the whole lung and GTV segmented.

**Figure 3.**
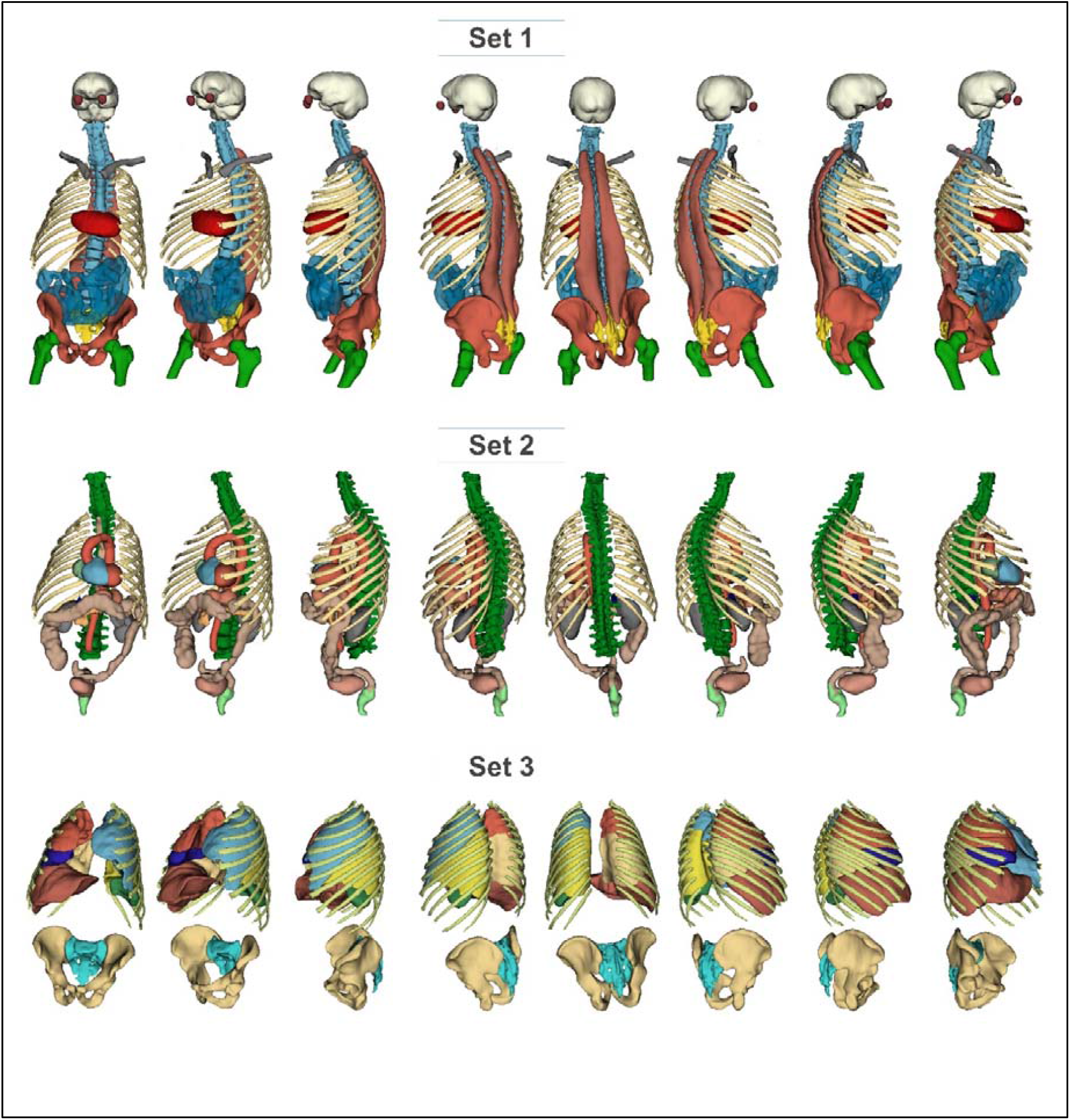
3D visualization of organ segmentations. Set1: Brain, eyeballs, vertebrae, clavicles, ribs, whole heart, rectus lumborum muscle, small intestine, sacrum, hips, and femoral heads. Set2: Vertebrae, oesophagus, aorta, heart substructures (LV, RV, LV cavity, RA, LA), stomach, pancreas, colon, rectum, and bladder. Set3: Lung five lobes, ribs, sacrum, hips. Some organs are repeated in all three sets for better visualization and as anatomical reference.

### Selected features

Table 4 shows the number of selected features for every 14 possible combinations of inputs where at least two types of inputs were used. In other words, CT GTV, PET GTV, CT Organomics, PET Organomics, and clinical parameters were not included in this Table since all the selected features were from the single input data. The most frequently selected features were for PET Organomics. The detailed names of features and organ names selected by all five FS models may be found in supplementary Table 2.

**Table 4.**
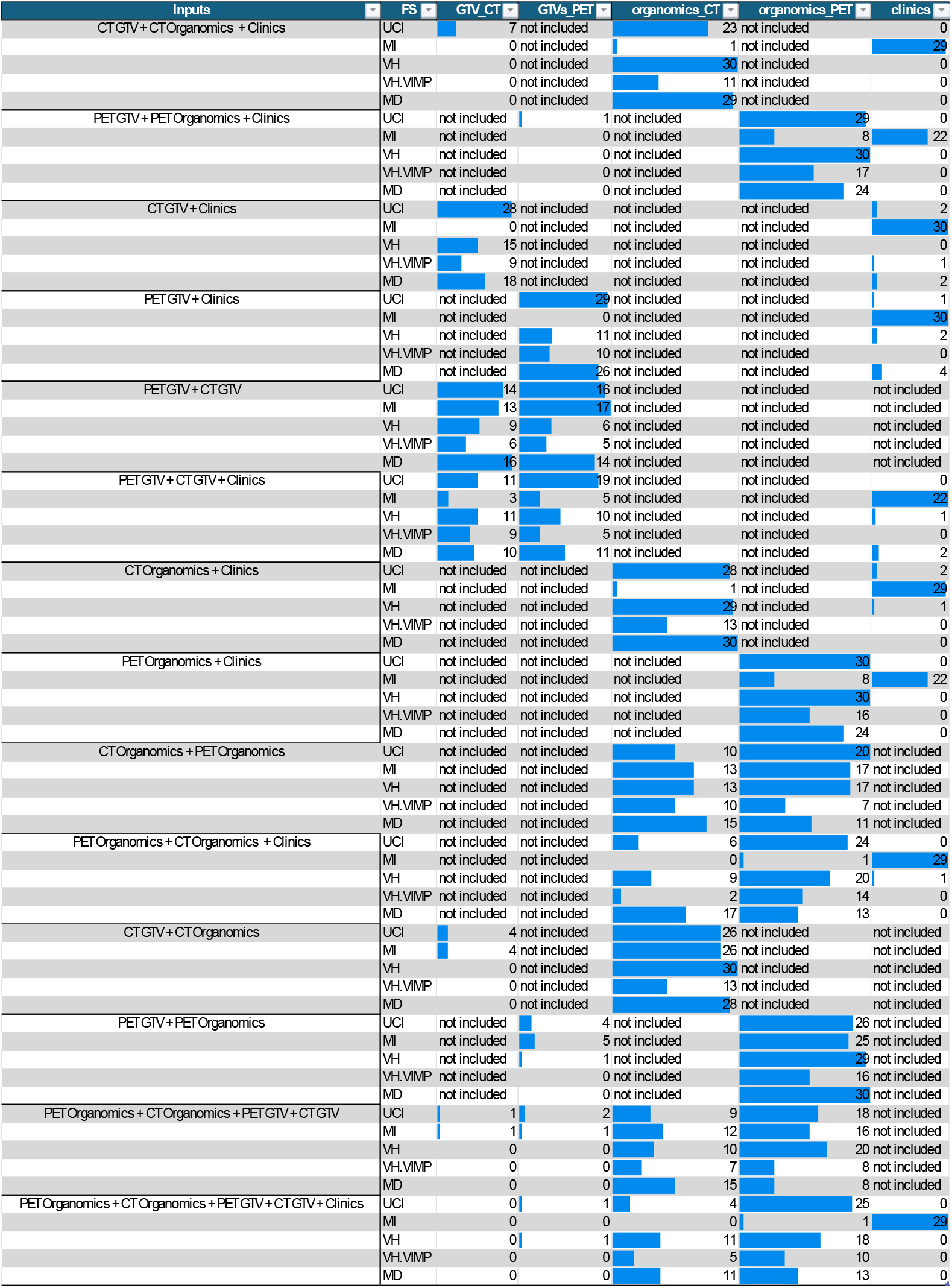
Frequency of selected features by every input data by all five feature selection methods. The blue color bar shows the frequency. In cases where input information was not used, the value was replaced by “not included”.

For inputs of PET Organomics + CT Organomics + PET GTV + CT GTV (all inputs except clinical information) all five feature selection methods selected mostly PET Organomics (86/150) and then CT Organomics (57/150) features, the most frequent selected organs by all FS methods were aorta, whole lung textures, heart left ventricle myocardium textures, and heart right ventricle textures.

### Model comparison

**Table *5*** summarizes the average and the best model C-Indices for every 19 combinations of inputs averaged over three folds. Supplementary Table 3 depicts the C-Index for every three folds and all 570 combinations of FS and models. The highest C-index (0.76) was achieved for a single fold using MD FS method, RSF machine and PET Organomics input. The resulting C-indices heatmap comparing all the 570 models are depicted in Figure 4.

**Table 5.** Model performance comparison based on inputs in terms of three-folds average C-Index. The highest values of 0.68 and 0.67 achieved are highlighted with bold font.

|  | # | Inputs | C-Index (average over all folds) |  |  |  |
| --- | --- | --- | --- | --- | --- | --- |
|  |  |  | mean | std | minimum | maximum |
| **Clinics Only** | **1** | Clinics | 0.58 | 0.02 | 0.52 | 0.61 |
| **GTV Only** | **2** | PET GTV | 0.59 | 0.02 | 0.55 | 0.63 |
|  | **3** | CT GTV | 0.59 | 0.02 | 0.55 | 0.62 |
|  | **4** | PET GTV + CT GTV | 0.59 | 0.02 | 0.55 | 0.63 |
|  | **5** | PET GTV + Clinics | 0.60 | 0.03 | 0.53 | 0.65 |
|  | **6** | CT GTV + Clinics | 0.59 | 0.02 | 0.54 | 0.63 |
|  | **7** | PET GTV + CT GTV + Clinics | 0.60 | 0.02 | 0.53 | 0.63 |
| **Organomics Only** | **8** | PET Organomics | 0.61 | 0.03 | 0.57 | **0.68** |
|  | **9** | CT Organomics | 0.61 | 0.02 | 0.55 | **0.67** |
|  | **10** | CT Organomics + PET Organomics | 0.60 | 0.03 | 0.52 | **0.68** |
|  | **11** | PET Organomics + Clinics | 0.60 | 0.02 | 0.57 | 0.65 |
|  | **12** | CT Organomics + Clinics | 0.60 | 0.02 | 0.57 | 0.63 |
|  | **13** | PET Organomics + CT Organomics + Clinics | 0.60 | 0.02 | 0.55 | 0.63 |
| **Single Modalities** | **14** | PET GTV + PET Organomics | 0.61 | 0.02 | 0.57 | 0.66 |
|  | **15** | CT GTV + CT Organomics | 0.60 | 0.02 | 0.56 | 0.65 |
|  | **16** | PET GTV + PET Organomics + Clinics | 0.60 | 0.03 | 0.54 | 0.66 |
|  | **17** | CT GTV + CT Organomics + Clinics | 0.59 | 0.02 | 0.52 | 0.62 |
|  | **18** | PET Organomics + CT Organomics + PET GTV + CT GTV | 0.59 | 0.02 | 0.55 | **0.67** |
| **All included** | **19** | PET Organomics + CT Organomics + PET GTV + CT GTV + Clinics | 0.60 | 0.02 | 0.55 | 0.65 |

**Figure 4.**
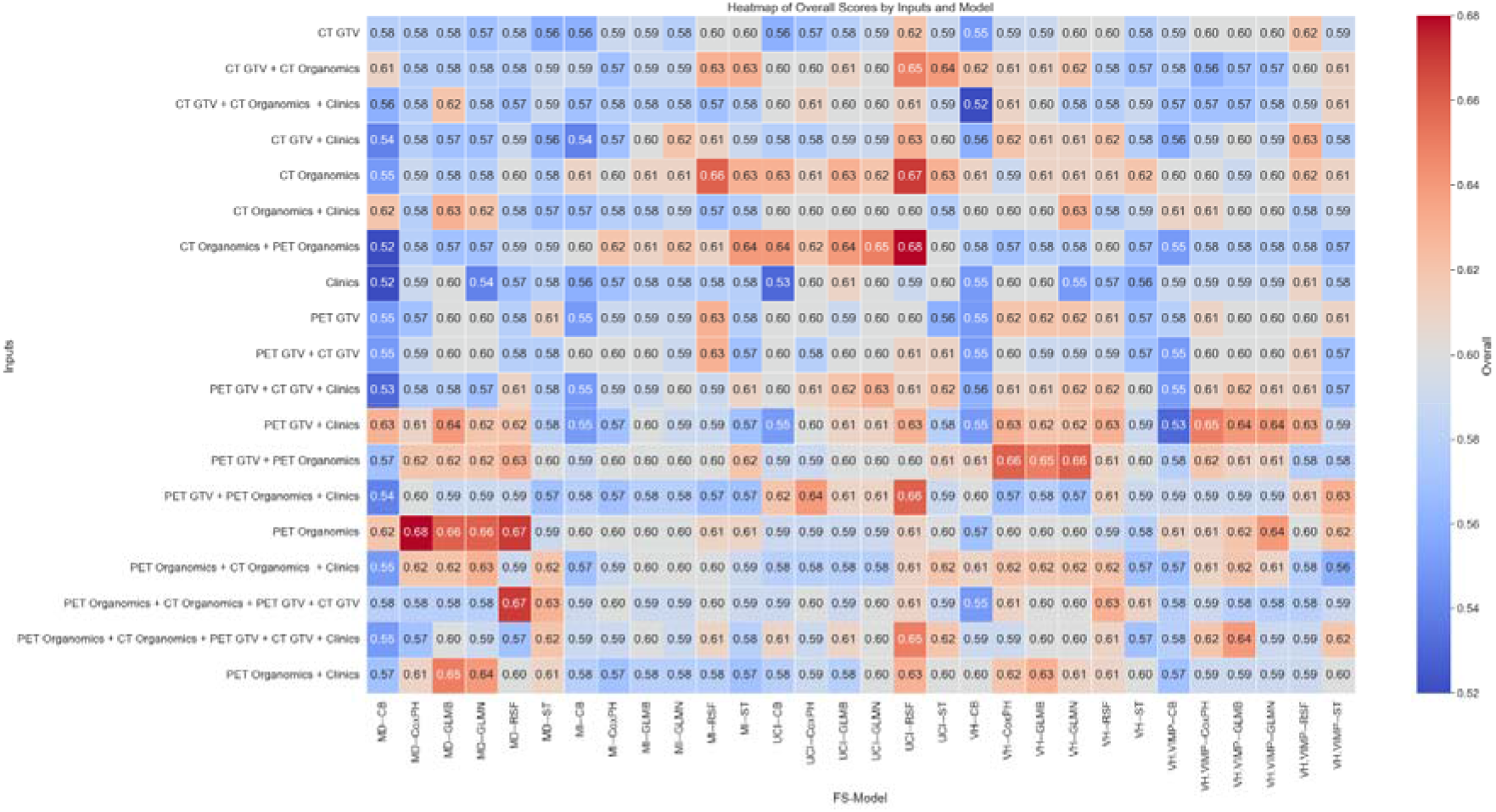
C-indices heatmap comparing all the 570 models. The colormap on the right shows the significance of colors. The vertical axis depicts the inputs whereas the horizontal access depicts the FS/Model combination.

Table 6 summarizes the inputs for every 30 combinations of FS and models with the highest C-Index averaged over all folds. PET Organomics was used as input in 18/30 of those combinations, while CT Organomics was used in 14/30 combinations. It should be noted that only 6/30 and 11/30 combinations used CT GTV and PET GTV radiomics.

**Table 6.**
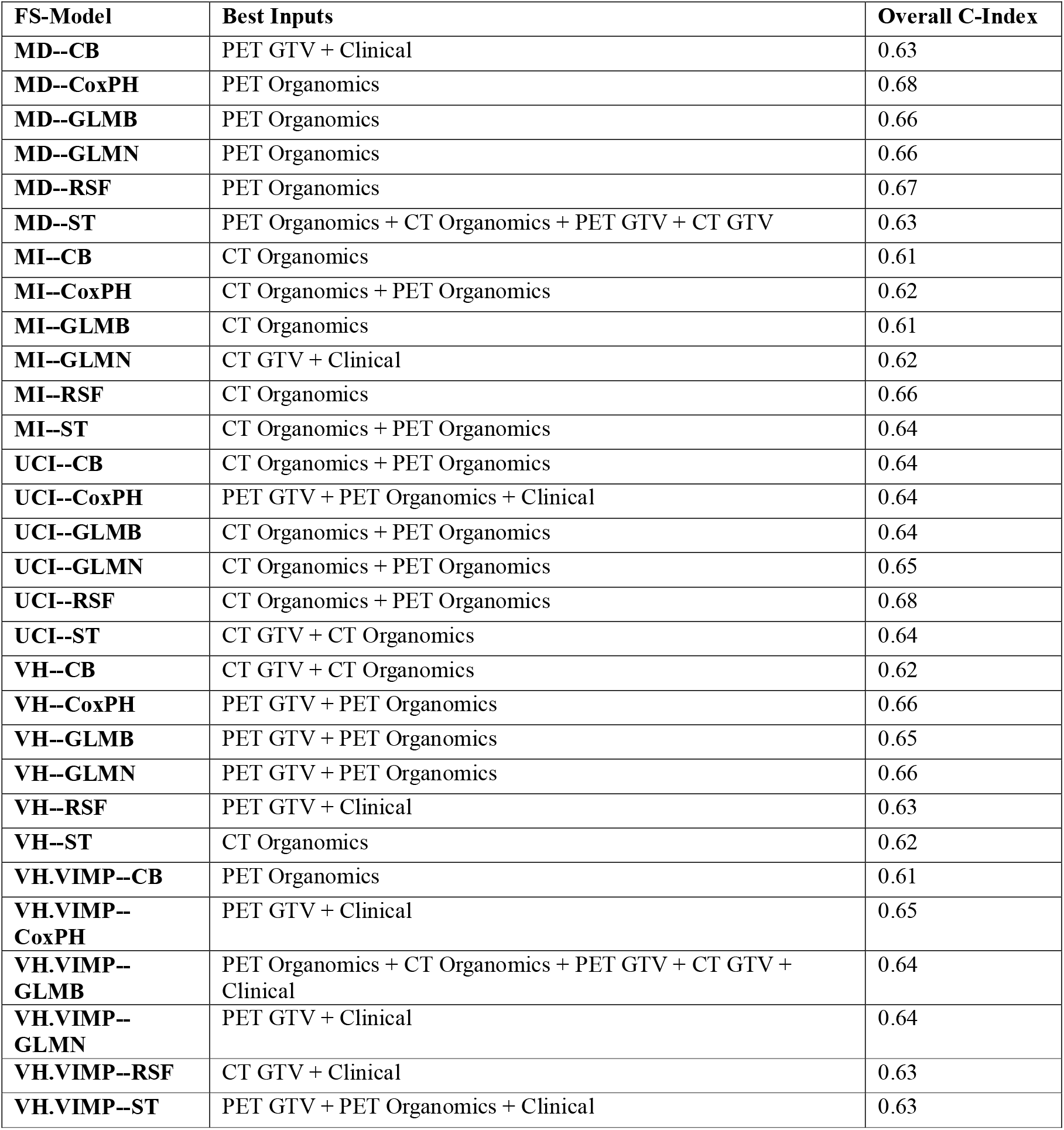
Highest C-Index and the corresponding inputs shown for every 30 combinations of FS/Model.

Figure 5 shows the Kaplan-Meier curves for nine selected models. GTV MD/RSF feature selection and model using PET Organomics + CT Organomics + PET GTV + CT as input showed the lowest p-value (0.00074), confirming its ability to separate high-risk patients from the low-risk group.

**Figure 5.**
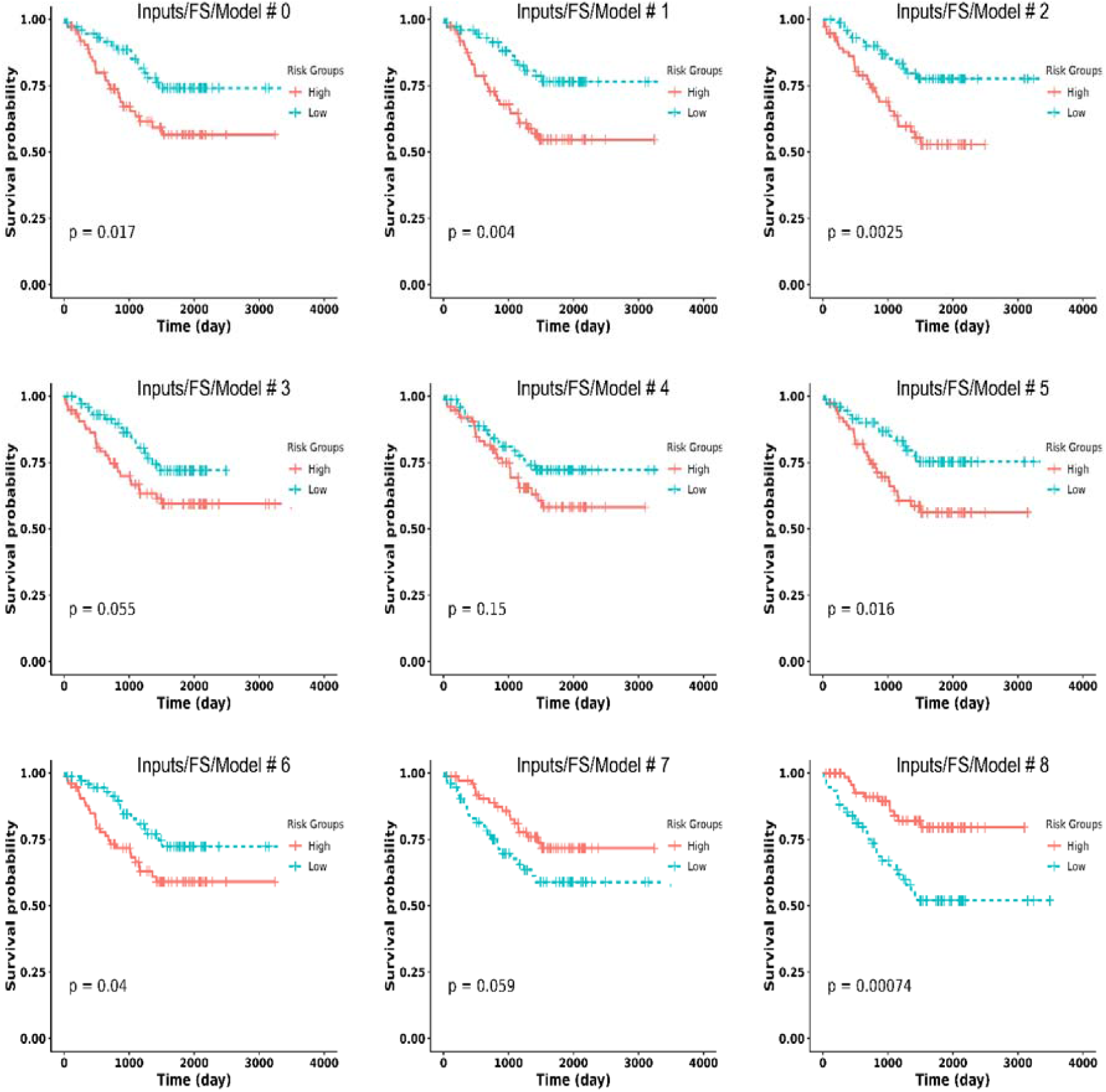
KM curves of 9 selected combinations of Inputs/FS/Model. #0: PET GTV + PET Organomics + Clinical/UCI/RSF, #1: PET Organomics + CT Organomics + PET GTV + CT GTV + Clinical/UCI/RSF, #2: CT Organomics/UCI/RSF, #3 CT Organomics/ MI/RSF, #4: PET Organomics/MD/Coxph, #5: CT Organomics + PET Organomics/UCI/glmnet, #6: CT GTV + CT Organomics/UCI/RSF, #7: CT Organomics + PET Organomics/VH/Coxph, #8: PET Organomics + CT Organomics + PET GTV + CT GTV/MD/RSF. P-values shown in the bottom of each curve. P-values <0.05 are considered statistically significant.

## Discussion

Survival prognosis information may be useful in optimizing treatment plans, risk stratification, and resource allocation. Artificial intelligence has proved promising in predicting the prognosis of patients with various malignancies [21; 22]. However, the potential information in regions other than the GTV is often overlooked and was not considered in NSCLC cancer prognosis. Lee et al. [6] used peri-tumoral regions radiomics and demonstrated its importance in two-year survival prediction. Hosny et al. [7] showed the importance of radiomics and dosiomics extracted from areas surrounding the GTV in NSCLC patients in prognosis through explainable deep learning and importance maps. Mattonen et al. [8] reported on the importance of metabolic tumour volume penumbra extended by 1cm in NSCLC recurrence. To the best of our knowledge, this is the first study exploring the added value of information contained in regions other than the treatment planning GTV and its surrounding tissues.

We explored the survival prediction capability of different sets of radiomic features extracted from different regions of the GTV and other organs from PET and CT imaging modalities. We also exploited the available clinical information and extensively tested 5 × 6 × 19 models in a three-fold data split to avoid the effect of random test/train split and invalid results. The aim of this study was to investigate the prognostic value of information extracted from different regions. Hence, we used multiple combinations of feature selection and machine learning methods to determine the approach achieving the best performance. Our results demonstrated that there is much more information in Organomics which can be used to predict the prognosis with AI. As summarized in Figure 4, all models achieving a C-Index more than 0.65 used Organomics, except one. The frequency of the selected features in Table 4 indicate the importance of Organomics in risk stratification, especially for the last two input combinations “PET Organomics + CT Organomics + PET GTV + CT GTV” and “PET Organomics + CT Organomics + PET GTV + CT GTV + Clinical” where the whole radiomics information from organs and GTVs was fed into feature selection algorithm and most of the selected features belong to PET Organomics and CT Organomics inputs for all FS methods, except for MI which selected clinical information instead and not the GTV information. Besides as presented in Table 6, most FS/Model combinations achieved the best results using PET Organomics and CT Organomics information. The most important organs affecting patients’ prognosis were the aorta, lungs, heart substructures.

Our best models using PET Organomics and CT Organomics C-Index averaged over three folds was 0.68, while the highest C-Index in a single fold was 0.76. Our best results using PET GTV, CT GTV and PET GTV + CT GTV in terms of C-Index were 0.63, 0.59, and 0.63, respectively which is in agreement with results reported by Amini et al. [5] using the same inputs (0.63, 0.64, and 0.65, respectively), except CT GTV where the C-Index achieved is lower in our study. It should be mentioned that we used three-fold cross-validation without harmonization w hile they used two-fold split strategy and ComBat harmonization. This comparison proved that although we did not have access to the manual GTV segmentations, our deep learning segmentation model provided a comparable GTV segmentation.

One limitation of our study was the lack of ground truth segmentation on PET/CT images. We tried to overcome this issue by using a large training dataset including the diagnostic CT for the same group of patients to train the state-of-the-art nnU-Net model through ensemble learning. We used CT images of PET/CT for the same group of patients as part of the training dataset. It should be clarified that the aim of this study was not to develop a generalizable deep learning segmentation model. This study aimed to test the hypothesis of the presence of important radiomics information in regions other than the GTV and its surrounding tissues. We used the deep learning models to transfer the segmentations from diagnostic CTs available in part of the dataset to PET/CT images. The overall Dice of 0.92 ± 0.08, actually comparable with results reported by Wang et al. [23] and Zhang et al. [24], demonstrated the successful transform of the segmentations. However, as we illustrate a case with Dice coefficient equal to 0.87, which is lower than average in Figure 2, there is a good match between the segmentations. We used two other datasets for training to overcome the image quality difference between diagnostic CT images and non-enhanced low-dose CT images of PET/CT. It should be mentioned that we cannot claim that organs other than the lungs were healthy organs, it may be additional pathologies in other areas which may be captured in the radiomics textures.

## Conclusion

There is important and useful information in terms of radiomic features outside the primary malignancies regions, including organs such as the aorta, heart, and lung which can improve the performance of AI algorithms. Our study suggests using as much as possible information from medical images toward generating a digital twin of patients with Organomics, GTV information and clinical data.

## Data Availability

Data is available online at: https://www.cancerimagingarchive.net/collection/nsclc-radiogenomics/

## Acknowledgements

This work was supported by the Euratom research and training programme 2019-2020 Sinfonia project under grant agreement No 945196.

## Compliance with ethical standards

This study was performed in line with the principles of the Declaration of Helsinki. Approval was granted by the local ethics committee. Consent forms were waived given the retrospective nature of the study.

