## Supplementary material for "Organomics: A concept reflecting the importance of PET/CT healthy organ radiomics in non-small cell lung cancer prognosis prediction using machine learning"

**Short running title:** Organomics: The importance of PET/CT healthy organ radiomics

- **Feature Selection explanation**

***Univariate C-index (UCI)***

In the (a) UCI method, the Spearman correlation test is performed at first and the redundant features ($R^2$>0.90) will be removed for further analysis. Thereafter, Concordance Index (C-index) is calculated for each feature by 100 bootstrapping in the training dataset. Subsequently, features are sorted by their mean C-indices, the top ten features are selected.

***Minimal Depth (MD), Variable hunting (VH), Variable hunting Variable Importance (VH.VIMP)***

MD, VH, and VH.VIMP are model-based FS methods that use Random Survival Forest (RSF) algorithm [1, 2]. MD sorts the features based on depth; the closer to the root node, the more predictive power is indicated. The top 10 features with MD are selected. In the VH method, the dataset is randomly split into training and testing datasets, followed by the application of RSF to the training dataset and the selection of random features via M-threshold. The initial model is created by selected features and adding features to the model will continue until the joint variable importance is stabilized. This procedure is repeated 50 times and features with the most frequency are selected. VH.VIMP functions similarly to the VH method except it uses variable importance instead of minimum depth to order the features, which is a faster method.

***Mutual Information (MI)***

MI has a completely parallelized performance. A linear approximation established on correlation is used to figure MI between two columns. Pearson's or Spearman's estimators are utilized to figure the correlation between continuous variables, whereas Somers' Dxy index is utilized to calculate the correlation between survival data [4].

- **Machine Learning models explanation**

***Cox Proportional Hazard regression (CoxPH):***
The hazard function is calculated by the Coxph model using the following equation:

$$h(t; X_1, X_2, \dots X_p) = h_0(t)\, \exp\,(\beta_1 X_1 + \cdots + \beta_p X_p)$$

wherein $h_0(t)$ represents the baseline hazard function, $X_j$ denotes the covariates (features), and $\beta_j$ denotes the coefficients [6]. The Cox partial likelihood function is maximized to approximate the vector of coefficients:

$$L(\beta) = \prod_{i=1\dots n\ S.t.\ \ \delta_i=1} \frac{\exp(\beta^T X^i)}{\sum_{l\in R_i} exp\ (\beta^T X^l)'}$$

***Cox Boosting (CB)**:*
Component-wise probability-based boosting is used to fit a Coxph model. The loss function for CB is CoxPH and partial likelihood estimation was used for penalizing the model in each boosting step [7].
***Generalized Linear Model Network (GLMN):***
Fits a penalized maximum likelihood model with a generalized linear model [8].
***GLM Boosting (GLMB):***
A boosting approach based on component-wise univariate linear models is used to fit a (generalized) linear model [9].
***Random Survival Forest (RSF):***
RSF is an ensemble tree algorithm developed to work on right-censored data. Breiman's random forest approach for survival data is extended in the Random Forests for Survival model [10].
***Survial Tree (ST):***

Essentially, ST combines a decision tree and a Cox method. These models can handle both time-to-event with right censor. A recursive partitioning approach, similar to that of decision trees, is used (8). Supplementary table 1 summarize the hyperparmerter that was optimized during optimization.

**Supplementary table 1.** Details of hyper-parameters optimization.

| **Model** | **R package** | **Hyperparameter** |
|---|---|---|
| Coxph | Survival | - |
| CB | CoxBoost | maxstepno : 50-500 |
| GLMN | glmnet | s: 0.001-0.1<br>alpha: 0-1 |
| RSF | randomForestSRC | split-rule: logrank, logrankscore<br>mtry: 1-10<br>nodesize: 1:20<br>ntree: 100, 500, 1000 |
| GLMB | mboost | mstop: 50-500 |
| ST | rpart | minsplit: 1-20<br>maxdepth: 1-30 |

Supplementary table 2 shows the list of selected features for five feature selection methods and inputs containing four PET GTV, CT GTV, PET Organomics, and CT Organomics.

**Supplementary table 2:** the detailed list of selected features selected by all five feature selection methods.

| **organs_plus_gtvgth_PET+CT_FN--Cindex** | **organs_plus_gtvgth_PET+CT_FN--MI** | **organs_plus_gtvgth_PET+CT_FN--VH** | **organs_plus_gtvgth_PET+CT_FN--VH.VIMP** | **organs_plus_gtvgth_PET+CT_FN--MD** |
|---|---|---|---|---|
| firstorder_RobustMeanAbsoluteDeviationBrain_PET_Label_1 | firstorder_InterquartileRangeLungs_binary_CT_Label_1 | glcm_IdmnRectusLumbarium_binary_PET_Label_1 | glszm_GrayLevelVarianceAorta_CT_Label_1 | glcm_IdmnVertebrae_binary_PET_Label_1 |
| glszm_SizeZoneNonUniformityNormalizedBrain_PET_Label_1 | glszm_SizeZoneNonUniformityNormalizedBrain_PET_Label_1 | glcm_IdmnVertebrae_binary_PET_Label_1 | firstorder_MedianHeartSubStructures_multi_CT_Label_3 | firstorder_MedianHeartSubStructures_multi_CT_Label_3 |
| glszm_SmallAreaLowGrayLevelEmphasisBrain_PET_Label_1 | firstorder_RobustMeanAbsoluteDeviationBrain_PET_Label_1 | gldm_DependenceNonUniformityAorta_PET_Label_1 | glszm_GrayLevelNonUniformityLungs_binary_PET_Label_1 | firstorder_MedianHeartSubStructures_multi_CT_Label_5 |
| gldm_DependenceEntropyLung_GTV_LIDC_NSCLC_PET_Label_1 | gldm_DependenceEntropyLung_GTV_LIDC_NSCLC_PET_Label_1 | gldm_SmallDependenceLowGrayLevelEmphasisHeartSubStructures_multi_PET_Label_5 | glcm_IdmnRibCage_binary_PET_Label_1 | firstorder_10PercentileHeartSubStructures_multi_CT_Label_3 |
| glrlm_GrayLevelNonUniformityLung_GTV_LIDC_NSCLC_CT_Label_1 | firstorder_10PercentileLungs_binary_CT_Label_1 | glrlm_LongRunHighGrayLevelEmphasisKidneys_binary_PET_Label_1 | glcm_IdmnVertebrae_binary_PET_Label_1 | gldm_DependenceEntropyAorta_CT_Label_1 |
| shape_FlatnessAdrenal_binary_CT_Label_1 | glszm_SmallAreaLowGrayLevelEmphasisBrain_PET_Label_1 | firstorder_SkewnessClavicles_binary_PET_Label_1 | ngtdm_ContrastAorta_PET_Label_1 | shape_MajorAxisLengthSpleen_CT_Label_1 |
| ngtdm_StrengthLungs_binary_PET_Label_1 | ngtdm_StrengthLungs_binary_PET_Label_1 | firstorder_MedianEsophagus_CT_Label_1 | gldm_SmallDependenceEmphasisClavicles_binary_PET_Label_1 | ngtdm_StrengthRectusLumbarium_binary_CT_Label_1 |
| glszm_SizeZoneNonUniformityLung_GTV_LIDC_NSCLC_PET_Label_1 | shape_FlatnessAdrenal_binary_CT_Label_1 | firstorder_MedianHeartSubStructures_multi_CT_Label_5 | glszm_SmallAreaLowGrayLevelEmphasisEsophagus_CT_Label_1 | glcm_MaximumProbabilityRibCage_binary_CT_Label_1 |
| gldm_SmallDependenceEmphasisClavicles_binary_PET_Label_1 | glrlm_GrayLevelNonUniformityLung_GTV_LIDC_NSCLC_CT_Label_1 | glszm_SizeZoneNonUniformityNormalizedHeartSubStructures_multi_PET_Label_2 | gldm_DependenceEntropyHeartSubStructures_multi_PET_Label_4 | glszm_ZoneEntropyStomach_PET_Label_1 |
| glszm_GrayLevelNonUniformityLu | glszm_GrayLevelNonUniformityLu | gldm_DependenceVarianceLungs_binary_PET_Label_1 | glrlm_RunEntropyHeartSubSt | firstorder_10PercentileHeartSubSt |

| | | | | |
|---|---|---|---|---|
| ngs_binary_PET_Label_1 | ngs_binary_PET_Label_1 | | ructures_multi_PET_Label_4 | ructures_multi_CT_Label_1 |
| ngtdm_ContrastAorta_PET_Label_1 | firstorder_InterquartileRangeLungs_binary_CT_Label_1 | ngtdm_StrengthClavicles_binary_PET_Label_1 | glrlm_GrayLevelNonUniformityAdrenal_binary_CT_Label_1 | glrlm_LongRunHighGrayLevelEmphasisEsophagus_CT_Label_1 |
| glcm_IdnAorta_PET_Label_1 | ngtdm_ContrastAorta_PET_Label_1 | firstorder_RangeRectusLumbarium_binary_CT_Label_1 | glcm_IdmnFemuralHeads_binary_CT_Label_1 | glszm_ZoneEntropyRectusLumbarium_binary_CT_Label_1 |
| glrlm_RunEntropyLungs_binary_PET_Label_1 | glcm_IdnAorta_PET_Label_1 | firstorder_MedianEsophagus_CT_Label_1 | firstorder_KurtosisHeartSubStructures_multi_CT_Label_2 | gldm_LargeDependenceHighGrayLevelEmphasisLungs_binary_CT_Label_1 |
| glrlm_RunEntropyLungs_binary_CT_Label_1 | glrlm_RunEntropyLungs_binary_PET_Label_1 | glszm_SmallAreaHighGrayLevelEmphasisStomach_PET_Label_1 | glrlm_ShortRunLowGrayLevelEmphasisStomach_PET_Label_1 | shape_Maximum3DDiameterUrinaryBladder_PET_Label_1 |
| glszm_GrayLevelNonUniformityLungs_binary_PET_Label_1 | glrlm_RunEntropyLungs_binary_CT_Label_1 | glcm_Imc2Aorta_CT_Label_1 | firstorder_SkewnessWholeHeart_CT_Label_1 | firstorder_InterquartileRangeLungs_binary_CT_Label_1 |
| glcm_IdmnAorta_PET_Label_1 | glszm_GrayLevelNonUniformityLungs_binary_PET_Label_1 | glszm_SmallAreaLowGrayLevelEmphasisEsophagus_CT_Label_1 | | glcm_IdmnFemuralHeads_binary_CT_Label_1 |
| glszm_SmallAreaLowGrayLevelEmphasisEsophagus_CT_Label_1 | glcm_IdmnAorta_PET_Label_1 | firstorder_KurtosisFemuralHeads_binary_CT_Label_1 | | shape_Maximum2DDiameterSliceHips_Sacrum_PET_Label_1 |
| firstorder_KurtosisEsophagus_PET_Label_1 | glszm_SmallAreaLowGrayLevelEmphasisEsophagus_CT_Label_1 | shape_MajorAxisLengthSpleen_PET_Label_1 | | firstorder_SkewnessWholeHeart_CT_Label_1 |
| ngtdm_StrengthLungs_binary_PET_Label_1 | ngtdm_StrengthLungs_binary_PET_Label_1 | glcm_DifferenceAverageUrinaryBladder_PET_Label_1 | | gldm_LowGrayLevelEmphasisStomach_PET_Label_1 |
| glrlm_LongRunLowGrayLevelEmphasisEsophagus_CT_Label_1 | glrlm_LongRunLowGrayLevelEmphasisEsophagus_CT_Label_1 | gldm_SmallDependenceHighGrayLevelEmphasisClavicles_binary_PET_Label_1 | | glrlm_ShortRunLowGrayLevelEmphasisStomach_PET_Label_1 |
| gldm_LargeDependenceHighGrayLevelEmphasisStomach_PET_Label_1 | firstorder_InterquartileRangeLungs_binary_CT_Label_1 | shape_FlatnessHeartSubStructures_multi_PET_Label_5 | | firstorder_MedianHeartSubStructures_multi_CT_Label_2 |
| glrlm_RunEntropyLungs_binary_CT_Label_1 | gldm_LargeDependenceHighGrayLevelEmphasisSt | ngtdm_StrengthHips_Sacrum_CT_Label_1 | | firstorder_KurtosisHeartSubStruct |

| | | | | |
|---|---|---|---|---|
| | omach_PET_Label_1 | | | ures_multi_PET_Label_5 |
| gldm_SmallDependenceLowGrayLevelEmphasisRibCage_binary_PET_Label_1 | glrlm_RunEntropyLungs_binary_CT_Label_1 | shape_Maximum2DDiameterSliceHips_Sacrum_PET_Label_1 | | glcm_MCCKidneys_binary_PET_Label_1 |
| shape_SphericitySpleen_CT_Label_1 | firstorder_MaximumVertebrae_binary_CT_Label_1 | glcm_MCCKidneys_binary_PET_Label_1 | | |
| glcm_CorrelationFemuralHeads_binary_CT_Label_1 | glszm_SizeZoneNonUniformityNormalizedBrain_PET_Label_1 | firstorder_10PercentileLungs_binary_CT_Label_1 | | |
| glrlm_RunLengthNonUniformityNormalizedBrain_PET_Label_1 | gldm_SmallDependenceLowGrayLevelEmphasisRibCage_binary_PET_Label_1 | ngtdm_BusynessAorta_CT_Label_1 | | |
| glszm_SizeZoneNonUniformityNormalizedBrain_PET_Label_1 | glrlm_RunLengthNonUniformityNormalizedBrain_PET_Label_1 | glszm_GrayLevelNonUniformityNormalizedClavicles_binary_PET_Label_1 | | |
| firstorder_SkewnessLungs_binary_PET_Label_1 | firstorder_SkewnessLungs_binary_PET_Label_1 | ngtdm_StrengthClavicles_binary_PET_Label_1 | | |
| glcm_Imc1Lungs_binary_CT_Label_1 | glcm_MCCLungs_binary_CT_Label_1 | glszm_GrayLevelNonUniformityEsophagus_PET_Label_1 | | |
| glcm_MCCLungs_binary_CT_Label_1 | glcm_CorrelationFemuralHeads_binary_CT_Label_1 | glszm_LowGrayLevelZoneEmphasisStomach_PET_Label_1 | | |

Supplementary Table 3 shows the detailed results of every folds and all combinations.

**Supplementary table 3:** the detailed results of performances in terms of C-Index for every three folds and all 570 combinations of inputs/FS/Models.

| Inputs | FS-Model | Fold1 | Fold2 | Fold3 | Overall |
|---|---|---|---|---|---|
| CT Organomics + PET Organomics | UCI--RSF | 0.71 | 0.62 | 0.7 | 0.68 |
| PET Organomics | MD--CoxPH | 0.74 | 0.64 | 0.66 | 0.68 |
| CT Organomics | UCI--RSF | 0.68 | 0.7 | 0.63 | 0.67 |
| PET Organomics | MD--RSF | 0.65 | 0.61 | 0.76 | 0.67 |
| PET Organomics + CT Organomics + PET GTV + CT GTV | MD--RSF | 0.7 | 0.56 | 0.75 | 0.67 |
| PET GTV + PET Organomics + Clinics | UCI--RSF | 0.62 | 0.66 | 0.69 | 0.66 |
| PET Organomics | MD--GLMN | 0.7 | 0.63 | 0.65 | 0.66 |

| | | | | | |
|---|---|---|---|---|---|
| PET Organomics | MD--GLMB | 0.7 | 0.63 | 0.65 | 0.66 |
| CT Organomics | MI--RSF | 0.66 | 0.69 | 0.62 | 0.66 |
| PET GTV + PET Organomics | VH--CoxPH | 0.59 | 0.68 | 0.7 | 0.66 |
| PET GTV + PET Organomics | VH--GLMN | 0.59 | 0.68 | 0.7 | 0.66 |
| PET Organomics + CT Organomics + PET GTV + CT GTV + Clinics | UCI--RSF | 0.6 | 0.69 | 0.64 | 0.65 |
| CT Organomics + PET Organomics | UCI--GLMN | 0.68 | 0.6 | 0.65 | 0.65 |
| CT GTV + CT Organomics | UCI--RSF | 0.65 | 0.7 | 0.6 | 0.65 |
| PET Organomics + Clinics | MD--GLMB | 0.66 | 0.59 | 0.71 | 0.65 |
| PET GTV + PET Organomics | VH--GLMB | 0.6 | 0.62 | 0.74 | 0.65 |
| PET GTV + Clinics | VH.VIMP--CoxPH | 0.58 | 0.68 | 0.67 | 0.65 |
| PET GTV + PET Organomics + Clinics | UCI--CoxPH | 0.59 | 0.65 | 0.68 | 0.64 |
| CT Organomics + PET Organomics | UCI--CB | 0.68 | 0.6 | 0.65 | 0.64 |
| CT Organomics + PET Organomics | UCI--GLMB | 0.7 | 0.58 | 0.65 | 0.64 |
| CT GTV + CT Organomics | UCI--ST | 0.66 | 0.66 | 0.61 | 0.64 |
| PET GTV + Clinics | MD--GLMB | 0.59 | 0.64 | 0.69 | 0.64 |
| PET Organomics + Clinics | MD--GLMN | 0.67 | 0.59 | 0.65 | 0.64 |
| CT Organomics + PET Organomics | MI--ST | 0.69 | 0.64 | 0.59 | 0.64 |
| PET Organomics + CT Organomics + PET GTV + CT GTV + Clinics | VH.VIMP--GLMB | 0.65 | 0.6 | 0.66 | 0.64 |
| PET GTV + Clinics | VH.VIMP--GLMN | 0.58 | 0.64 | 0.69 | 0.64 |
| PET GTV + Clinics | VH.VIMP--GLMB | 0.58 | 0.66 | 0.69 | 0.64 |
| PET Organomics | VH.VIMP--GLMN | 0.67 | 0.55 | 0.69 | 0.64 |
| CT GTV + Clinics | UCI--RSF | 0.59 | 0.65 | 0.63 | 0.63 |
| PET GTV + Clinics | UCI--RSF | 0.59 | 0.68 | 0.6 | 0.63 |
| PET GTV + CT GTV + Clinics | UCI--GLMN | 0.59 | 0.7 | 0.6 | 0.63 |
| CT Organomics | UCI--CB | 0.66 | 0.6 | 0.62 | 0.63 |
| CT Organomics | UCI--GLMB | 0.66 | 0.6 | 0.62 | 0.63 |
| CT Organomics | UCI--ST | 0.65 | 0.64 | 0.6 | 0.63 |
| PET Organomics + Clinics | UCI--RSF | 0.58 | 0.64 | 0.67 | 0.63 |
| PET GTV + Clinics | MD--CB | 0.59 | 0.64 | 0.66 | 0.63 |
| CT Organomics + Clinics | MD--GLMB | 0.62 | 0.56 | 0.69 | 0.63 |
| PET Organomics + CT Organomics + Clinics | MD--GLMN | 0.58 | 0.6 | 0.71 | 0.63 |
| PET GTV + PET Organomics | MD--RSF | 0.59 | 0.57 | 0.75 | 0.63 |
| PET Organomics + CT Organomics + PET GTV + CT GTV | MD--ST | 0.58 | 0.67 | 0.64 | 0.63 |
| PET GTV | MI--RSF | 0.59 | 0.66 | 0.64 | 0.63 |
| PET GTV + CT GTV | MI--RSF | 0.59 | 0.71 | 0.59 | 0.63 |
| CT Organomics | MI--ST | 0.58 | 0.69 | 0.62 | 0.63 |
| CT GTV + CT Organomics | MI--RSF | 0.64 | 0.65 | 0.59 | 0.63 |
| CT GTV + CT Organomics | MI--ST | 0.61 | 0.67 | 0.61 | 0.63 |
| PET GTV + Clinics | VH--CoxPH | 0.59 | 0.72 | 0.59 | 0.63 |
| PET GTV + Clinics | VH--RSF | 0.6 | 0.65 | 0.63 | 0.63 |
| CT Organomics + Clinics | VH--GLMN | 0.65 | 0.56 | 0.68 | 0.63 |

| | | | | | |
|---|---|---|---|---|---|
| PET Organomics + Clinics | VH--GLMB | 0.64 | 0.65 | 0.6 | 0.63 |
| PET Organomics + CT Organomics + PET GTV + CT GTV | VH--RSF | 0.65 | 0.62 | 0.63 | 0.63 |
| PET GTV + PET Organomics + Clinics | VH.VIMP--ST | 0.69 | 0.56 | 0.65 | 0.63 |
| CT GTV + Clinics | VH.VIMP--RSF | 0.61 | 0.69 | 0.59 | 0.63 |
| PET GTV + Clinics | VH.VIMP--RSF | 0.59 | 0.61 | 0.69 | 0.63 |
| PET GTV + PET Organomics + Clinics | UCI--CB | 0.59 | 0.61 | 0.65 | 0.62 |
| PET Organomics + CT Organomics + PET GTV + CT GTV + Clinics | UCI--ST | 0.58 | 0.55 | 0.73 | 0.62 |
| CT GTV | UCI--RSF | 0.6 | 0.66 | 0.59 | 0.62 |
| PET GTV + CT GTV + Clinics | UCI--GLMB | 0.59 | 0.66 | 0.61 | 0.62 |
| PET GTV + CT GTV + Clinics | UCI--ST | 0.62 | 0.66 | 0.57 | 0.62 |
| CT Organomics | UCI--GLMN | 0.67 | 0.59 | 0.61 | 0.62 |
| CT Organomics + PET Organomics | UCI--CoxPH | 0.66 | 0.61 | 0.6 | 0.62 |
| PET Organomics + CT Organomics + Clinics | UCI--ST | 0.59 | 0.58 | 0.68 | 0.62 |
| CT GTV + CT Organomics + Clinics | MD--GLMB | 0.63 | 0.57 | 0.68 | 0.62 |
| PET Organomics + CT Organomics + PET GTV + CT GTV + Clinics | MD--ST | 0.61 | 0.6 | 0.66 | 0.62 |
| PET GTV + Clinics | MD--GLMN | 0.59 | 0.63 | 0.63 | 0.62 |
| PET GTV + Clinics | MD--RSF | 0.6 | 0.67 | 0.59 | 0.62 |
| PET Organomics | MD--CB | 0.71 | 0.5 | 0.65 | 0.62 |
| CT Organomics + Clinics | MD--CB | 0.62 | 0.56 | 0.67 | 0.62 |
| CT Organomics + Clinics | MD--GLMN | 0.62 | 0.56 | 0.68 | 0.62 |
| PET Organomics + CT Organomics + Clinics | MD--CoxPH | 0.58 | 0.57 | 0.72 | 0.62 |
| PET Organomics + CT Organomics + Clinics | MD--GLMB | 0.58 | 0.6 | 0.69 | 0.62 |
| PET Organomics + CT Organomics + Clinics | MD--ST | 0.59 | 0.59 | 0.68 | 0.62 |
| PET GTV + PET Organomics | MD--CoxPH | 0.59 | 0.63 | 0.65 | 0.62 |
| PET GTV + PET Organomics | MD--GLMN | 0.58 | 0.62 | 0.64 | 0.62 |
| PET GTV + PET Organomics | MD--GLMB | 0.58 | 0.63 | 0.64 | 0.62 |
| CT GTV + Clinics | MI--GLMN | 0.62 | 0.56 | 0.68 | 0.62 |
| CT Organomics + PET Organomics | MI--CoxPH | 0.67 | 0.6 | 0.57 | 0.62 |
| CT Organomics + PET Organomics | MI--GLMN | 0.69 | 0.59 | 0.57 | 0.62 |
| PET GTV + PET Organomics | MI--ST | 0.65 | 0.6 | 0.61 | 0.62 |
| PET GTV | VH--CoxPH | 0.61 | 0.66 | 0.58 | 0.62 |
| PET GTV | VH--GLMN | 0.62 | 0.66 | 0.57 | 0.62 |
| PET GTV | VH--GLMB | 0.61 | 0.66 | 0.57 | 0.62 |
| CT GTV + Clinics | VH--CoxPH | 0.58 | 0.69 | 0.61 | 0.62 |
| CT GTV + Clinics | VH--RSF | 0.59 | 0.69 | 0.59 | 0.62 |
| PET GTV + Clinics | VH--GLMN | 0.59 | 0.69 | 0.57 | 0.62 |
| PET GTV + Clinics | VH--GLMB | 0.59 | 0.69 | 0.58 | 0.62 |
| PET GTV + CT GTV + Clinics | VH--GLMN | 0.61 | 0.69 | 0.57 | 0.62 |
| PET GTV + CT GTV + Clinics | VH--RSF | 0.6 | 0.67 | 0.58 | 0.62 |
| CT Organomics | VH--ST | 0.64 | 0.53 | 0.7 | 0.62 |
| PET Organomics + Clinics | VH--CoxPH | 0.64 | 0.64 | 0.57 | 0.62 |
| PET Organomics + CT Organomics + Clinics | VH--CoxPH | 0.61 | 0.57 | 0.67 | 0.62 |

| | | | | | |
|---|---|---|---|---|---|
| PET Organomics + CT Organomics + Clinics | VH--GLMN | 0.61 | 0.58 | 0.67 | 0.62 |
| PET Organomics + CT Organomics + Clinics | VH--RSF | 0.66 | 0.56 | 0.63 | 0.62 |
| PET Organomics + CT Organomics + Clinics | VH--GLMB | 0.61 | 0.57 | 0.69 | 0.62 |
| CT GTV + CT Organomics | VH--CB | 0.71 | 0.56 | 0.59 | 0.62 |
| CT GTV + CT Organomics | VH--GLMN | 0.72 | 0.56 | 0.59 | 0.62 |
| PET Organomics + CT Organomics + PET GTV + CT GTV + Clinics | VH.VIMP--CoxPH | 0.63 | 0.58 | 0.65 | 0.62 |
| PET Organomics + CT Organomics + PET GTV + CT GTV + Clinics | VH.VIMP--ST | 0.63 | 0.57 | 0.65 | 0.62 |
| CT GTV | VH.VIMP--RSF | 0.6 | 0.62 | 0.64 | 0.62 |
| PET GTV + CT GTV + Clinics | VH.VIMP--GLMB | 0.59 | 0.69 | 0.57 | 0.62 |
| CT Organomics | VH.VIMP--RSF | 0.61 | 0.68 | 0.58 | 0.62 |
| PET Organomics | VH.VIMP--GLMB | 0.64 | 0.55 | 0.65 | 0.62 |
| PET Organomics | VH.VIMP--ST | 0.69 | 0.55 | 0.62 | 0.62 |
| PET Organomics + CT Organomics + Clinics | VH.VIMP--GLMB | 0.63 | 0.57 | 0.66 | 0.62 |
| PET GTV + PET Organomics | VH.VIMP--CoxPH | 0.7 | 0.56 | 0.6 | 0.62 |
| CT GTV + CT Organomics + Clinics | UCI--CoxPH | 0.62 | 0.6 | 0.6 | 0.61 |
| CT GTV + CT Organomics + Clinics | UCI--RSF | 0.58 | 0.62 | 0.62 | 0.61 |
| PET GTV + PET Organomics + Clinics | UCI--GLMN | 0.6 | 0.58 | 0.65 | 0.61 |
| PET GTV + PET Organomics + Clinics | UCI--GLMB | 0.59 | 0.59 | 0.65 | 0.61 |
| PET Organomics + CT Organomics + PET GTV + CT GTV + Clinics | UCI--CB | 0.62 | 0.6 | 0.61 | 0.61 |
| PET Organomics + CT Organomics + PET GTV + CT GTV + Clinics | UCI--GLMB | 0.62 | 0.59 | 0.6 | 0.61 |
| Clinics | UCI--GLMB | 0.58 | 0.61 | 0.65 | 0.61 |
| PET GTV + CT GTV | UCI--RSF | 0.59 | 0.66 | 0.57 | 0.61 |
| PET GTV + CT GTV | UCI--ST | 0.58 | 0.62 | 0.64 | 0.61 |
| PET GTV + Clinics | UCI--GLMN | 0.59 | 0.63 | 0.61 | 0.61 |
| PET GTV + Clinics | UCI--GLMB | 0.59 | 0.64 | 0.6 | 0.61 |
| PET GTV + CT GTV + Clinics | UCI--CoxPH | 0.59 | 0.64 | 0.58 | 0.61 |
| PET GTV + CT GTV + Clinics | UCI--RSF | 0.59 | 0.66 | 0.58 | 0.61 |
| CT Organomics | UCI--CoxPH | 0.63 | 0.6 | 0.61 | 0.61 |
| PET Organomics | UCI--RSF | 0.65 | 0.61 | 0.58 | 0.61 |
| PET Organomics + CT Organomics + Clinics | UCI--RSF | 0.6 | 0.65 | 0.59 | 0.61 |
| CT GTV + CT Organomics | UCI--GLMB | 0.61 | 0.6 | 0.61 | 0.61 |
| PET GTV + PET Organomics | UCI--ST | 0.67 | 0.57 | 0.61 | 0.61 |
| PET Organomics + CT Organomics + PET GTV + CT GTV | UCI--RSF | 0.61 | 0.62 | 0.6 | 0.61 |
| PET GTV | MD--ST | 0.68 | 0.58 | 0.57 | 0.61 |
| PET GTV + Clinics | MD--CoxPH | 0.59 | 0.62 | 0.64 | 0.61 |
| PET GTV + CT GTV + Clinics | MD--RSF | 0.59 | 0.68 | 0.58 | 0.61 |
| PET Organomics + Clinics | MD--CoxPH | 0.62 | 0.59 | 0.63 | 0.61 |

| | | | | | |
|---|---|---|---|---|---|
| PET Organomics + Clinics | MD--ST | 0.62 | 0.6 | 0.6 | 0.61 |
| CT GTV + CT Organomics | MD--CB | 0.59 | 0.57 | 0.67 | 0.61 |
| PET Organomics + CT Organomics + PET GTV + CT GTV + Clinics | MI--RSF | 0.57 | 0.59 | 0.66 | 0.61 |
| CT GTV + Clinics | MI--RSF | 0.62 | 0.56 | 0.65 | 0.61 |
| PET GTV + CT GTV + Clinics | MI--ST | 0.61 | 0.66 | 0.56 | 0.61 |
| CT Organomics | MI--CB | 0.65 | 0.6 | 0.58 | 0.61 |
| CT Organomics | MI--GLMN | 0.66 | 0.59 | 0.58 | 0.61 |
| CT Organomics | MI--GLMB | 0.66 | 0.59 | 0.58 | 0.61 |
| PET Organomics | MI--RSF | 0.67 | 0.6 | 0.57 | 0.61 |
| PET Organomics | MI--ST | 0.66 | 0.57 | 0.59 | 0.61 |
| CT Organomics + PET Organomics | MI--RSF | 0.66 | 0.6 | 0.58 | 0.61 |
| CT Organomics + PET Organomics | MI--GLMB | 0.67 | 0.59 | 0.57 | 0.61 |
| CT GTV + CT Organomics + Clinics | VH--CoxPH | 0.66 | 0.58 | 0.57 | 0.61 |
| PET GTV + PET Organomics + Clinics | VH--RSF | 0.57 | 0.63 | 0.63 | 0.61 |
| PET Organomics + CT Organomics + PET GTV + CT GTV + Clinics | VH--RSF | 0.58 | 0.56 | 0.67 | 0.61 |
| PET GTV | VH--RSF | 0.6 | 0.63 | 0.61 | 0.61 |
| CT GTV + Clinics | VH--GLMN | 0.58 | 0.68 | 0.58 | 0.61 |
| CT GTV + Clinics | VH--GLMB | 0.58 | 0.66 | 0.59 | 0.61 |
| PET GTV + CT GTV + Clinics | VH--CoxPH | 0.6 | 0.64 | 0.58 | 0.61 |
| PET GTV + CT GTV + Clinics | VH--GLMB | 0.6 | 0.66 | 0.57 | 0.61 |
| CT Organomics | VH--CB | 0.67 | 0.58 | 0.58 | 0.61 |
| CT Organomics | VH--GLMN | 0.67 | 0.59 | 0.58 | 0.61 |
| CT Organomics | VH--RSF | 0.65 | 0.57 | 0.6 | 0.61 |
| CT Organomics | VH--GLMB | 0.67 | 0.59 | 0.57 | 0.61 |
| PET Organomics + Clinics | VH--GLMN | 0.64 | 0.64 | 0.57 | 0.61 |
| PET Organomics + Clinics | VH--RSF | 0.65 | 0.59 | 0.57 | 0.61 |
| PET Organomics + CT Organomics + Clinics | VH--CB | 0.66 | 0.58 | 0.59 | 0.61 |
| CT GTV + CT Organomics | VH--CoxPH | 0.72 | 0.56 | 0.57 | 0.61 |
| CT GTV + CT Organomics | VH--GLMB | 0.69 | 0.56 | 0.59 | 0.61 |
| PET GTV + PET Organomics | VH--CB | 0.59 | 0.5 | 0.74 | 0.61 |
| PET GTV + PET Organomics | VH--RSF | 0.6 | 0.6 | 0.63 | 0.61 |
| PET Organomics + CT Organomics + PET GTV + CT GTV | VH--CoxPH | 0.65 | 0.6 | 0.59 | 0.61 |
| PET Organomics + CT Organomics + PET GTV + CT GTV | VH--ST | 0.58 | 0.63 | 0.63 | 0.61 |
| CT GTV + CT Organomics + Clinics | VH.VIMP--ST | 0.58 | 0.56 | 0.68 | 0.61 |
| PET GTV + PET Organomics + Clinics | VH.VIMP--RSF | 0.66 | 0.57 | 0.59 | 0.61 |
| Clinics | VH.VIMP--RSF | 0.6 | 0.57 | 0.66 | 0.61 |
| PET GTV | VH.VIMP--CoxPH | 0.61 | 0.66 | 0.57 | 0.61 |
| PET GTV | VH.VIMP--ST | 0.58 | 0.64 | 0.61 | 0.61 |
| PET GTV + CT GTV | VH.VIMP--RSF | 0.6 | 0.59 | 0.64 | 0.61 |

| | | | | | |
|---|---|---|---|---|---|
| PET GTV + CT GTV + Clinics | VH.VIMP--CoxPH | 0.59 | 0.67 | 0.58 | 0.61 |
| PET GTV + CT GTV + Clinics | VH.VIMP--GLMN | 0.59 | 0.65 | 0.57 | 0.61 |
| PET GTV + CT GTV + Clinics | VH.VIMP--RSF | 0.62 | 0.64 | 0.58 | 0.61 |
| CT Organomics | VH.VIMP--ST | 0.56 | 0.67 | 0.6 | 0.61 |
| PET Organomics | VH.VIMP--CoxPH | 0.64 | 0.56 | 0.63 | 0.61 |
| PET Organomics | VH.VIMP--CB | 0.59 | 0.56 | 0.69 | 0.61 |
| CT Organomics + Clinics | VH.VIMP--CoxPH | 0.59 | 0.57 | 0.67 | 0.61 |
| CT Organomics + Clinics | VH.VIMP--CB | 0.58 | 0.57 | 0.67 | 0.61 |
| PET Organomics + CT Organomics + Clinics | VH.VIMP--CoxPH | 0.61 | 0.58 | 0.62 | 0.61 |
| PET Organomics + CT Organomics + Clinics | VH.VIMP--GLMN | 0.63 | 0.57 | 0.62 | 0.61 |
| CT GTV + CT Organomics | VH.VIMP--ST | 0.58 | 0.56 | 0.68 | 0.61 |
| PET GTV + PET Organomics | VH.VIMP--GLMN | 0.71 | 0.56 | 0.58 | 0.61 |
| PET GTV + PET Organomics | VH.VIMP--GLMB | 0.69 | 0.56 | 0.58 | 0.61 |
| CT GTV + CT Organomics + Clinics | UCI--CB | 0.57 | 0.63 | 0.6 | 0.6 |
| CT GTV + CT Organomics + Clinics | UCI--GLMN | 0.57 | 0.63 | 0.61 | 0.6 |
| CT GTV + CT Organomics + Clinics | UCI--GLMB | 0.57 | 0.64 | 0.6 | 0.6 |
| PET Organomics + CT Organomics + PET GTV + CT GTV + Clinics | UCI--GLMN | 0.61 | 0.59 | 0.59 | 0.6 |
| Clinics | UCI--CoxPH | 0.57 | 0.62 | 0.62 | 0.6 |
| Clinics | UCI--GLMN | 0.58 | 0.57 | 0.66 | 0.6 |
| Clinics | UCI--ST | 0.57 | 0.63 | 0.6 | 0.6 |
| PET GTV | UCI--CoxPH | 0.59 | 0.62 | 0.6 | 0.6 |
| PET GTV | UCI--CB | 0.59 | 0.64 | 0.57 | 0.6 |
| PET GTV | UCI--GLMN | 0.58 | 0.64 | 0.58 | 0.6 |
| PET GTV | UCI--RSF | 0.6 | 0.62 | 0.57 | 0.6 |
| PET GTV + CT GTV | UCI--CB | 0.58 | 0.63 | 0.58 | 0.6 |
| PET GTV + CT GTV | UCI--GLMN | 0.58 | 0.64 | 0.59 | 0.6 |
| PET GTV + CT GTV | UCI--GLMB | 0.58 | 0.64 | 0.58 | 0.6 |
| CT GTV + Clinics | UCI--ST | 0.61 | 0.63 | 0.55 | 0.6 |
| PET GTV + Clinics | UCI--CoxPH | 0.6 | 0.63 | 0.57 | 0.6 |
| PET GTV + CT GTV + Clinics | UCI--CB | 0.59 | 0.64 | 0.57 | 0.6 |
| PET Organomics | UCI--ST | 0.64 | 0.58 | 0.57 | 0.6 |
| CT Organomics + PET Organomics | UCI--ST | 0.61 | 0.61 | 0.58 | 0.6 |
| CT Organomics + Clinics | UCI--CoxPH | 0.57 | 0.62 | 0.6 | 0.6 |
| CT Organomics + Clinics | UCI--CB | 0.57 | 0.63 | 0.6 | 0.6 |
| CT Organomics + Clinics | UCI--GLMN | 0.57 | 0.62 | 0.6 | 0.6 |
| CT Organomics + Clinics | UCI--RSF | 0.59 | 0.63 | 0.57 | 0.6 |

| | | | | | |
|---|---|---|---|---|---|
| CT Organomics + Clinics | UCI--GLMB | 0.57 | 0.62 | 0.6 | 0.6 |
| PET Organomics + Clinics | UCI--GLMN | 0.58 | 0.57 | 0.63 | 0.6 |
| PET Organomics + Clinics | UCI--ST | 0.59 | 0.61 | 0.6 | 0.6 |
| CT GTV + CT Organomics | UCI--CoxPH | 0.6 | 0.59 | 0.61 | 0.6 |
| CT GTV + CT Organomics | UCI--CB | 0.62 | 0.56 | 0.62 | 0.6 |
| CT GTV + CT Organomics | UCI--GLMN | 0.6 | 0.61 | 0.6 | 0.6 |
| PET GTV + PET Organomics | UCI--GLMN | 0.61 | 0.59 | 0.59 | 0.6 |
| PET GTV + PET Organomics | UCI--RSF | 0.64 | 0.61 | 0.56 | 0.6 |
| PET GTV + PET Organomics | UCI--GLMB | 0.61 | 0.59 | 0.59 | 0.6 |
| PET Organomics + CT Organomics + PET GTV + CT GTV | UCI--CoxPH | 0.6 | 0.61 | 0.58 | 0.6 |
| PET Organomics + CT Organomics + PET GTV + CT GTV | UCI--GLMN | 0.61 | 0.6 | 0.6 | 0.6 |
| PET GTV + PET Organomics + Clinics | MD--CoxPH | 0.58 | 0.6 | 0.62 | 0.6 |
| PET Organomics + CT Organomics + PET GTV + CT GTV + Clinics | MD--GLMB | 0.58 | 0.6 | 0.62 | 0.6 |
| Clinics | MD--GLMB | 0.57 | 0.59 | 0.64 | 0.6 |
| PET GTV | MD--GLMN | 0.58 | 0.65 | 0.57 | 0.6 |
| PET GTV | MD--GLMB | 0.58 | 0.66 | 0.56 | 0.6 |
| PET GTV + CT GTV | MD--GLMN | 0.57 | 0.64 | 0.57 | 0.6 |
| PET GTV + CT GTV | MD--GLMB | 0.57 | 0.63 | 0.59 | 0.6 |
| CT Organomics | MD--RSF | 0.62 | 0.55 | 0.61 | 0.6 |
| PET Organomics + Clinics | MD--RSF | 0.64 | 0.57 | 0.57 | 0.6 |
| PET GTV + PET Organomics | MD--ST | 0.6 | 0.61 | 0.58 | 0.6 |
| PET Organomics + CT Organomics + PET GTV + CT GTV + Clinics | MI--GLMB | 0.58 | 0.58 | 0.66 | 0.6 |
| CT GTV | MI--RSF | 0.6 | 0.63 | 0.58 | 0.6 |
| CT GTV | MI--ST | 0.59 | 0.57 | 0.65 | 0.6 |
| PET GTV + CT GTV | MI--CoxPH | 0.58 | 0.6 | 0.6 | 0.6 |
| PET GTV + CT GTV | MI--CB | 0.58 | 0.63 | 0.59 | 0.6 |
| PET GTV + CT GTV | MI--GLMB | 0.58 | 0.62 | 0.59 | 0.6 |
| CT GTV + Clinics | MI--GLMB | 0.56 | 0.56 | 0.68 | 0.6 |
| PET GTV + Clinics | MI--GLMB | 0.56 | 0.56 | 0.66 | 0.6 |
| PET GTV + CT GTV + Clinics | MI--GLMN | 0.58 | 0.6 | 0.61 | 0.6 |
| CT Organomics | MI--CoxPH | 0.65 | 0.56 | 0.58 | 0.6 |
| PET Organomics | MI--CoxPH | 0.64 | 0.59 | 0.58 | 0.6 |
| PET Organomics | MI--CB | 0.64 | 0.58 | 0.58 | 0.6 |
| PET Organomics | MI--GLMN | 0.64 | 0.59 | 0.58 | 0.6 |
| PET Organomics | MI--GLMB | 0.63 | 0.58 | 0.58 | 0.6 |
| CT Organomics + PET Organomics | MI--CB | 0.63 | 0.59 | 0.57 | 0.6 |
| PET Organomics + CT Organomics + Clinics | MI--GLMN | 0.6 | 0.58 | 0.63 | 0.6 |
| PET Organomics + CT Organomics + Clinics | MI--RSF | 0.58 | 0.58 | 0.66 | 0.6 |
| PET Organomics + CT Organomics + Clinics | MI--GLMB | 0.57 | 0.57 | 0.64 | 0.6 |
| PET GTV + PET Organomics | MI--CoxPH | 0.62 | 0.59 | 0.59 | 0.6 |
| PET GTV + PET Organomics | MI--GLMN | 0.62 | 0.59 | 0.59 | 0.6 |
| PET GTV + PET Organomics | MI--RSF | 0.63 | 0.62 | 0.56 | 0.6 |
| PET GTV + PET Organomics | MI--GLMB | 0.62 | 0.58 | 0.59 | 0.6 |

| | | | | | |
|---|---|---|---|---|---|
| PET Organomics + CT Organomics + PET GTV + CT GTV | MI--CoxPH | 0.6 | 0.61 | 0.58 | 0.6 |
| PET Organomics + CT Organomics + PET GTV + CT GTV | MI--RSF | 0.61 | 0.6 | 0.58 | 0.6 |
| CT GTV + CT Organomics + Clinics | VH--GLMB | 0.65 | 0.57 | 0.58 | 0.6 |
| PET GTV + PET Organomics + Clinics | VH--CB | 0.63 | 0.56 | 0.61 | 0.6 |
| PET Organomics + CT Organomics + PET GTV + CT GTV + Clinics | VH--GLMN | 0.61 | 0.6 | 0.58 | 0.6 |
| PET Organomics + CT Organomics + PET GTV + CT GTV + Clinics | VH--GLMB | 0.6 | 0.6 | 0.6 | 0.6 |
| Clinics | VH--CoxPH | 0.59 | 0.59 | 0.63 | 0.6 |
| Clinics | VH--GLMB | 0.59 | 0.58 | 0.64 | 0.6 |
| CT GTV | VH--GLMN | 0.58 | 0.62 | 0.58 | 0.6 |
| CT GTV | VH--RSF | 0.58 | 0.63 | 0.58 | 0.6 |
| PET GTV + CT GTV | VH--CoxPH | 0.58 | 0.63 | 0.57 | 0.6 |
| PET GTV + CT GTV + Clinics | VH--ST | 0.59 | 0.64 | 0.58 | 0.6 |
| PET Organomics | VH--CoxPH | 0.58 | 0.64 | 0.58 | 0.6 |
| PET Organomics | VH--GLMN | 0.58 | 0.63 | 0.58 | 0.6 |
| PET Organomics | VH--GLMB | 0.57 | 0.59 | 0.62 | 0.6 |
| CT Organomics + PET Organomics | VH--RSF | 0.6 | 0.62 | 0.59 | 0.6 |
| CT Organomics + Clinics | VH--CoxPH | 0.67 | 0.56 | 0.57 | 0.6 |
| CT Organomics + Clinics | VH--CB | 0.57 | 0.56 | 0.68 | 0.6 |
| CT Organomics + Clinics | VH--GLMB | 0.57 | 0.56 | 0.66 | 0.6 |
| PET Organomics + Clinics | VH--CB | 0.63 | 0.58 | 0.6 | 0.6 |
| PET Organomics + Clinics | VH--ST | 0.62 | 0.63 | 0.57 | 0.6 |
| PET GTV + PET Organomics | VH--ST | 0.56 | 0.66 | 0.57 | 0.6 |
| PET Organomics + CT Organomics + PET GTV + CT GTV | VH--GLMN | 0.66 | 0.56 | 0.59 | 0.6 |
| PET Organomics + CT Organomics + PET GTV + CT GTV | VH--GLMB | 0.65 | 0.56 | 0.59 | 0.6 |
| CT GTV | VH.VIMP--CoxPH | 0.6 | 0.62 | 0.57 | 0.6 |
| CT GTV | VH.VIMP--GLMN | 0.6 | 0.62 | 0.58 | 0.6 |
| CT GTV | VH.VIMP--GLMB | 0.6 | 0.62 | 0.57 | 0.6 |
| PET GTV | VH.VIMP--GLMN | 0.59 | 0.66 | 0.57 | 0.6 |
| PET GTV | VH.VIMP--RSF | 0.6 | 0.62 | 0.59 | 0.6 |
| PET GTV | VH.VIMP--GLMB | 0.59 | 0.64 | 0.57 | 0.6 |
| PET GTV + CT GTV | VH.VIMP--CoxPH | 0.58 | 0.63 | 0.58 | 0.6 |
| PET GTV + CT GTV | VH.VIMP--GLMN | 0.58 | 0.62 | 0.58 | 0.6 |
| PET GTV + CT GTV | VH.VIMP--GLMB | 0.58 | 0.63 | 0.58 | 0.6 |
| CT GTV + Clinics | VH.VIMP--GLMB | 0.58 | 0.62 | 0.59 | 0.6 |

| | | | | | |
|---|---|---|---|---|---|
| CT Organomics | VH.VIMP--CoxPH | 0.63 | 0.56 | 0.59 | 0.6 |
| CT Organomics | VH.VIMP--CB | 0.63 | 0.57 | 0.61 | 0.6 |
| CT Organomics | VH.VIMP--GLMN | 0.63 | 0.57 | 0.59 | 0.6 |
| PET Organomics | VH.VIMP--RSF | 0.6 | 0.56 | 0.63 | 0.6 |
| CT Organomics + Clinics | VH.VIMP--GLMN | 0.59 | 0.57 | 0.66 | 0.6 |
| CT Organomics + Clinics | VH.VIMP--GLMB | 0.59 | 0.57 | 0.65 | 0.6 |
| PET Organomics + Clinics | VH.VIMP--ST | 0.67 | 0.56 | 0.57 | 0.6 |
| CT GTV + CT Organomics | VH.VIMP--RSF | 0.67 | 0.57 | 0.57 | 0.6 |
| CT GTV + CT Organomics + Clinics | UCI--ST | 0.62 | 0.57 | 0.59 | 0.59 |
| PET GTV + PET Organomics + Clinics | UCI--ST | 0.6 | 0.6 | 0.58 | 0.59 |
| PET Organomics + CT Organomics + PET GTV + CT GTV + Clinics | UCI--CoxPH | 0.6 | 0.57 | 0.59 | 0.59 |
| Clinics | UCI--RSF | 0.58 | 0.6 | 0.6 | 0.59 |
| CT GTV | UCI--GLMN | 0.58 | 0.58 | 0.59 | 0.59 |
| CT GTV | UCI--ST | 0.57 | 0.63 | 0.56 | 0.59 |
| PET GTV | UCI--GLMB | 0.59 | 0.63 | 0.57 | 0.59 |
| CT GTV + Clinics | UCI--GLMN | 0.59 | 0.61 | 0.57 | 0.59 |
| CT GTV + Clinics | UCI--GLMB | 0.59 | 0.59 | 0.57 | 0.59 |
| PET Organomics | UCI--CoxPH | 0.6 | 0.59 | 0.58 | 0.59 |
| PET Organomics | UCI--CB | 0.61 | 0.59 | 0.58 | 0.59 |
| PET Organomics | UCI--GLMN | 0.61 | 0.59 | 0.59 | 0.59 |
| PET Organomics | UCI--GLMB | 0.6 | 0.58 | 0.59 | 0.59 |
| PET Organomics + Clinics | UCI--CoxPH | 0.59 | 0.56 | 0.63 | 0.59 |
| PET GTV + PET Organomics | UCI--CoxPH | 0.61 | 0.58 | 0.59 | 0.59 |
| PET GTV + PET Organomics | UCI--CB | 0.61 | 0.59 | 0.57 | 0.59 |
| PET Organomics + CT Organomics + PET GTV + CT GTV | UCI--CB | 0.6 | 0.6 | 0.57 | 0.59 |
| PET Organomics + CT Organomics + PET GTV + CT GTV | UCI--GLMB | 0.6 | 0.6 | 0.57 | 0.59 |
| PET Organomics + CT Organomics + PET GTV + CT GTV | UCI--ST | 0.59 | 0.6 | 0.58 | 0.59 |
| CT GTV + CT Organomics + Clinics | MD--ST | 0.6 | 0.6 | 0.58 | 0.59 |
| PET GTV + PET Organomics + Clinics | MD--GLMN | 0.58 | 0.55 | 0.64 | 0.59 |
| PET GTV + PET Organomics + Clinics | MD--RSF | 0.61 | 0.57 | 0.59 | 0.59 |
| PET GTV + PET Organomics + Clinics | MD--GLMB | 0.58 | 0.55 | 0.64 | 0.59 |
| PET Organomics + CT Organomics + PET GTV + CT GTV + Clinics | MD--GLMN | 0.58 | 0.58 | 0.61 | 0.59 |
| Clinics | MD--CoxPH | 0.57 | 0.58 | 0.63 | 0.59 |
| PET GTV + CT GTV | MD--CoxPH | 0.58 | 0.64 | 0.57 | 0.59 |
| CT GTV + Clinics | MD--RSF | 0.6 | 0.6 | 0.58 | 0.59 |
| CT Organomics | MD--CoxPH | 0.6 | 0.56 | 0.6 | 0.59 |
| PET Organomics | MD--ST | 0.59 | 0.59 | 0.57 | 0.59 |

| | | | | | |
|---|---|---|---|---|---|
| CT Organomics + PET Organomics | MD--RSF | 0.59 | 0.57 | 0.59 | 0.59 |
| CT Organomics + PET Organomics | MD--ST | 0.58 | 0.67 | 0.53 | 0.59 |
| PET Organomics + CT Organomics + Clinics | MD--RSF | 0.58 | 0.62 | 0.57 | 0.59 |
| CT GTV + CT Organomics | MD--ST | 0.61 | 0.56 | 0.6 | 0.59 |
| PET Organomics + CT Organomics + PET GTV + CT GTV + Clinics | MI--CoxPH | 0.59 | 0.57 | 0.6 | 0.59 |
| PET Organomics + CT Organomics + PET GTV + CT GTV + Clinics | MI--CB | 0.57 | 0.56 | 0.65 | 0.59 |
| PET Organomics + CT Organomics + PET GTV + CT GTV + Clinics | MI--GLMN | 0.57 | 0.57 | 0.63 | 0.59 |
| CT GTV | MI--CoxPH | 0.59 | 0.56 | 0.62 | 0.59 |
| CT GTV | MI--GLMB | 0.59 | 0.58 | 0.59 | 0.59 |
| PET GTV | MI--CoxPH | 0.58 | 0.59 | 0.6 | 0.59 |
| PET GTV | MI--GLMN | 0.58 | 0.64 | 0.56 | 0.59 |
| PET GTV | MI--GLMB | 0.59 | 0.62 | 0.57 | 0.59 |
| PET GTV + CT GTV | MI--GLMN | 0.58 | 0.62 | 0.59 | 0.59 |
| CT GTV + Clinics | MI--ST | 0.61 | 0.56 | 0.58 | 0.59 |
| PET GTV + Clinics | MI--GLMN | 0.62 | 0.56 | 0.59 | 0.59 |
| PET GTV + Clinics | MI--RSF | 0.62 | 0.56 | 0.58 | 0.59 |
| PET GTV + CT GTV + Clinics | MI--CoxPH | 0.57 | 0.61 | 0.58 | 0.59 |
| PET GTV + CT GTV + Clinics | MI--RSF | 0.57 | 0.62 | 0.59 | 0.59 |
| PET GTV + CT GTV + Clinics | MI--GLMB | 0.58 | 0.6 | 0.58 | 0.59 |
| CT Organomics + Clinics | MI--GLMN | 0.58 | 0.57 | 0.61 | 0.59 |
| PET Organomics + CT Organomics + Clinics | MI--CoxPH | 0.59 | 0.57 | 0.6 | 0.59 |
| PET Organomics + CT Organomics + Clinics | MI--ST | 0.57 | 0.57 | 0.63 | 0.59 |
| CT GTV + CT Organomics | MI--CB | 0.6 | 0.6 | 0.58 | 0.59 |
| CT GTV + CT Organomics | MI--GLMN | 0.59 | 0.59 | 0.58 | 0.59 |
| CT GTV + CT Organomics | MI--GLMB | 0.59 | 0.59 | 0.58 | 0.59 |
| PET GTV + PET Organomics | MI--CB | 0.6 | 0.59 | 0.58 | 0.59 |
| PET Organomics + CT Organomics + PET GTV + CT GTV | MI--CB | 0.61 | 0.58 | 0.57 | 0.59 |
| PET Organomics + CT Organomics + PET GTV + CT GTV | MI--GLMN | 0.6 | 0.6 | 0.57 | 0.59 |
| PET Organomics + CT Organomics + PET GTV + CT GTV | MI--GLMB | 0.6 | 0.59 | 0.57 | 0.59 |
| PET Organomics + CT Organomics + PET GTV + CT GTV | MI--ST | 0.59 | 0.61 | 0.59 | 0.59 |
| CT GTV + CT Organomics + Clinics | VH--ST | 0.64 | 0.55 | 0.58 | 0.59 |
| PET GTV + PET Organomics + Clinics | VH--ST | 0.59 | 0.55 | 0.64 | 0.59 |
| PET Organomics + CT Organomics + PET GTV + CT GTV + Clinics | VH--CoxPH | 0.6 | 0.59 | 0.59 | 0.59 |
| PET Organomics + CT Organomics + PET GTV + CT GTV + Clinics | VH--CB | 0.6 | 0.6 | 0.56 | 0.59 |
| CT GTV | VH--CoxPH | 0.58 | 0.62 | 0.58 | 0.59 |
| CT GTV | VH--GLMB | 0.58 | 0.62 | 0.58 | 0.59 |
| PET GTV + CT GTV | VH--GLMN | 0.58 | 0.62 | 0.57 | 0.59 |
| PET GTV + CT GTV | VH--RSF | 0.59 | 0.6 | 0.57 | 0.59 |
| PET GTV + CT GTV | VH--GLMB | 0.58 | 0.63 | 0.57 | 0.59 |
| PET GTV + Clinics | VH--ST | 0.59 | 0.61 | 0.58 | 0.59 |

| | | | | | |
|---|---|---|---|---|---|
| CT Organomics | VH--CoxPH | 0.6 | 0.58 | 0.57 | 0.59 |
| PET Organomics | VH--RSF | 0.61 | 0.59 | 0.57 | 0.59 |
| CT Organomics + Clinics | VH--ST | 0.58 | 0.56 | 0.63 | 0.59 |
| CT GTV + CT Organomics + Clinics | VH.VIMP--RSF | 0.6 | 0.56 | 0.62 | 0.59 |
| PET GTV + PET Organomics + Clinics | VH.VIMP--CoxPH | 0.63 | 0.57 | 0.57 | 0.59 |
| PET GTV + PET Organomics + Clinics | VH.VIMP--CB | 0.65 | 0.57 | 0.56 | 0.59 |
| PET GTV + PET Organomics + Clinics | VH.VIMP--GLMN | 0.65 | 0.57 | 0.57 | 0.59 |
| PET GTV + PET Organomics + Clinics | VH.VIMP--GLMB | 0.65 | 0.56 | 0.56 | 0.59 |
| PET Organomics + CT Organomics + PET GTV + CT GTV + Clinics | VH.VIMP--GLMN | 0.67 | 0.59 | 0.5 | 0.59 |
| PET Organomics + CT Organomics + PET GTV + CT GTV + Clinics | VH.VIMP--RSF | 0.62 | 0.57 | 0.58 | 0.59 |
| Clinics | VH.VIMP--CoxPH | 0.58 | 0.56 | 0.62 | 0.59 |
| Clinics | VH.VIMP--CB | 0.58 | 0.57 | 0.62 | 0.59 |
| Clinics | VH.VIMP--GLMN | 0.58 | 0.56 | 0.62 | 0.59 |
| Clinics | VH.VIMP--GLMB | 0.58 | 0.56 | 0.62 | 0.59 |
| CT GTV | VH.VIMP--CB | 0.6 | 0.61 | 0.57 | 0.59 |
| CT GTV | VH.VIMP--ST | 0.57 | 0.64 | 0.57 | 0.59 |
| CT GTV + Clinics | VH.VIMP--CoxPH | 0.58 | 0.62 | 0.59 | 0.59 |
| CT GTV + Clinics | VH.VIMP--GLMN | 0.58 | 0.61 | 0.59 | 0.59 |
| PET GTV + Clinics | VH.VIMP--ST | 0.59 | 0.58 | 0.6 | 0.59 |
| CT Organomics | VH.VIMP--GLMB | 0.63 | 0.56 | 0.59 | 0.59 |
| CT Organomics + Clinics | VH.VIMP--ST | 0.59 | 0.6 | 0.59 | 0.59 |
| PET Organomics + Clinics | VH.VIMP--CoxPH | 0.64 | 0.56 | 0.57 | 0.59 |
| PET Organomics + Clinics | VH.VIMP--GLMN | 0.64 | 0.56 | 0.57 | 0.59 |
| PET Organomics + Clinics | VH.VIMP--RSF | 0.59 | 0.6 | 0.58 | 0.59 |
| PET Organomics + Clinics | VH.VIMP--GLMB | 0.64 | 0.56 | 0.57 | 0.59 |
| PET Organomics + CT Organomics + PET GTV + CT GTV | VH.VIMP--CoxPH | 0.59 | 0.56 | 0.61 | 0.59 |
| PET Organomics + CT Organomics + PET GTV + CT GTV | VH.VIMP--ST | 0.58 | 0.58 | 0.6 | 0.59 |
| CT GTV | UCI--GLMB | 0.58 | 0.57 | 0.59 | 0.58 |
| PET GTV + CT GTV | UCI--CoxPH | 0.58 | 0.57 | 0.58 | 0.58 |

| | | | | | |
|---|---|---|---|---|---|
| CT GTV + Clinics | UCI--CoxPH | 0.6 | 0.57 | 0.58 | 0.58 |
| CT GTV + Clinics | UCI--CB | 0.59 | 0.57 | 0.57 | 0.58 |
| PET GTV + Clinics | UCI--ST | 0.57 | 0.61 | 0.56 | 0.58 |
| CT Organomics + Clinics | UCI--ST | 0.58 | 0.59 | 0.57 | 0.58 |
| PET Organomics + Clinics | UCI--CB | 0.58 | 0.57 | 0.6 | 0.58 |
| PET Organomics + Clinics | UCI--GLMB | 0.59 | 0.57 | 0.6 | 0.58 |
| PET Organomics + CT Organomics + Clinics | UCI--CoxPH | 0.58 | 0.6 | 0.57 | 0.58 |
| PET Organomics + CT Organomics + Clinics | UCI--CB | 0.57 | 0.6 | 0.58 | 0.58 |
| PET Organomics + CT Organomics + Clinics | UCI--GLMN | 0.57 | 0.6 | 0.57 | 0.58 |
| PET Organomics + CT Organomics + Clinics | UCI--GLMB | 0.58 | 0.6 | 0.58 | 0.58 |
| CT GTV + CT Organomics + Clinics | MD--CoxPH | 0.62 | 0.57 | 0.57 | 0.58 |
| CT GTV + CT Organomics + Clinics | MD--GLMN | 0.62 | 0.57 | 0.57 | 0.58 |
| Clinics | MD--ST | 0.58 | 0.59 | 0.55 | 0.58 |
| CT GTV | MD--CoxPH | 0.6 | 0.56 | 0.58 | 0.58 |
| CT GTV | MD--CB | 0.59 | 0.58 | 0.59 | 0.58 |
| CT GTV | MD--RSF | 0.58 | 0.58 | 0.59 | 0.58 |
| CT GTV | MD--GLMB | 0.59 | 0.56 | 0.59 | 0.58 |
| PET GTV | MD--RSF | 0.59 | 0.57 | 0.57 | 0.58 |
| PET GTV + CT GTV | MD--RSF | 0.56 | 0.6 | 0.59 | 0.58 |
| PET GTV + CT GTV | MD--ST | 0.58 | 0.6 | 0.56 | 0.58 |
| CT GTV + Clinics | MD--CoxPH | 0.59 | 0.56 | 0.57 | 0.58 |
| PET GTV + Clinics | MD--ST | 0.57 | 0.6 | 0.57 | 0.58 |
| PET GTV + CT GTV + Clinics | MD--CoxPH | 0.59 | 0.59 | 0.57 | 0.58 |
| PET GTV + CT GTV + Clinics | MD--GLMB | 0.58 | 0.59 | 0.57 | 0.58 |
| PET GTV + CT GTV + Clinics | MD--ST | 0.6 | 0.55 | 0.59 | 0.58 |
| CT Organomics | MD--GLMN | 0.59 | 0.56 | 0.58 | 0.58 |
| CT Organomics | MD--GLMB | 0.59 | 0.56 | 0.59 | 0.58 |
| CT Organomics | MD--ST | 0.57 | 0.58 | 0.58 | 0.58 |
| CT Organomics + PET Organomics | MD--CoxPH | 0.57 | 0.57 | 0.58 | 0.58 |
| CT Organomics + Clinics | MD--CoxPH | 0.62 | 0.56 | 0.56 | 0.58 |
| CT Organomics + Clinics | MD--RSF | 0.6 | 0.57 | 0.56 | 0.58 |
| CT GTV + CT Organomics | MD--CoxPH | 0.59 | 0.57 | 0.59 | 0.58 |
| CT GTV + CT Organomics | MD--GLMN | 0.59 | 0.56 | 0.58 | 0.58 |
| CT GTV + CT Organomics | MD--RSF | 0.6 | 0.56 | 0.57 | 0.58 |
| CT GTV + CT Organomics | MD--GLMB | 0.58 | 0.57 | 0.58 | 0.58 |
| PET Organomics + CT Organomics + PET GTV + CT GTV | MD--CoxPH | 0.59 | 0.56 | 0.6 | 0.58 |
| PET Organomics + CT Organomics + PET GTV + CT GTV | MD--CB | 0.6 | 0.56 | 0.59 | 0.58 |
| PET Organomics + CT Organomics + PET GTV + CT GTV | MD--GLMN | 0.6 | 0.56 | 0.58 | 0.58 |
| PET Organomics + CT Organomics + PET GTV + CT GTV | MD--GLMB | 0.6 | 0.56 | 0.59 | 0.58 |
| CT GTV + CT Organomics + Clinics | MI--CoxPH | 0.59 | 0.57 | 0.59 | 0.58 |
| CT GTV + CT Organomics + Clinics | MI--GLMN | 0.6 | 0.58 | 0.58 | 0.58 |
| CT GTV + CT Organomics + Clinics | MI--GLMB | 0.58 | 0.58 | 0.58 | 0.58 |
| CT GTV + CT Organomics + Clinics | MI--ST | 0.57 | 0.6 | 0.57 | 0.58 |

| | | | | | |
|---|---|---|---|---|---|
| PET GTV + PET Organomics + Clinics | MI--CB | 0.59 | 0.58 | 0.57 | 0.58 |
| PET GTV + PET Organomics + Clinics | MI--GLMN | 0.59 | 0.59 | 0.57 | 0.58 |
| PET GTV + PET Organomics + Clinics | MI--GLMB | 0.59 | 0.59 | 0.57 | 0.58 |
| PET Organomics + CT Organomics + PET GTV + CT GTV + Clinics | MI--ST | 0.57 | 0.57 | 0.61 | 0.58 |
| Clinics | MI--GLMN | 0.59 | 0.57 | 0.6 | 0.58 |
| Clinics | MI--RSF | 0.57 | 0.56 | 0.6 | 0.58 |
| Clinics | MI--GLMB | 0.56 | 0.57 | 0.62 | 0.58 |
| Clinics | MI--ST | 0.61 | 0.55 | 0.59 | 0.58 |
| CT GTV | MI--GLMN | 0.59 | 0.56 | 0.59 | 0.58 |
| PET GTV | MI--ST | 0.59 | 0.59 | 0.57 | 0.58 |
| CT Organomics + Clinics | MI--CoxPH | 0.59 | 0.57 | 0.58 | 0.58 |
| CT Organomics + Clinics | MI--GLMB | 0.58 | 0.58 | 0.57 | 0.58 |
| CT Organomics + Clinics | MI--ST | 0.6 | 0.57 | 0.57 | 0.58 |
| PET Organomics + Clinics | MI--CB | 0.58 | 0.57 | 0.57 | 0.58 |
| PET Organomics + Clinics | MI--GLMN | 0.58 | 0.58 | 0.57 | 0.58 |
| PET Organomics + Clinics | MI--RSF | 0.58 | 0.57 | 0.59 | 0.58 |
| PET Organomics + Clinics | MI--GLMB | 0.58 | 0.58 | 0.57 | 0.58 |
| CT GTV + CT Organomics + Clinics | VH--GLMN | 0.66 | 0.57 | 0.5 | 0.58 |
| CT GTV + CT Organomics + Clinics | VH--RSF | 0.62 | 0.56 | 0.57 | 0.58 |
| PET GTV + PET Organomics + Clinics | VH--GLMB | 0.58 | 0.56 | 0.61 | 0.58 |
| CT GTV | VH--ST | 0.57 | 0.6 | 0.57 | 0.58 |
| CT GTV + Clinics | VH--ST | 0.59 | 0.56 | 0.59 | 0.58 |
| PET Organomics | VH--ST | 0.64 | 0.58 | 0.53 | 0.58 |
| CT Organomics + PET Organomics | VH--CB | 0.61 | 0.57 | 0.57 | 0.58 |
| CT Organomics + PET Organomics | VH--GLMN | 0.61 | 0.57 | 0.56 | 0.58 |
| CT Organomics + PET Organomics | VH--GLMB | 0.61 | 0.56 | 0.56 | 0.58 |
| CT Organomics + Clinics | VH--RSF | 0.57 | 0.57 | 0.59 | 0.58 |
| CT GTV + CT Organomics | VH--RSF | 0.6 | 0.56 | 0.57 | 0.58 |
| CT GTV + CT Organomics + Clinics | VH.VIMP--GLMN | 0.57 | 0.56 | 0.59 | 0.58 |
| PET Organomics + CT Organomics + PET GTV + CT GTV + Clinics | VH.VIMP--CB | 0.65 | 0.59 | 0.5 | 0.58 |
| Clinics | VH.VIMP--ST | 0.58 | 0.57 | 0.58 | 0.58 |
| PET GTV | VH.VIMP--CB | 0.59 | 0.6 | 0.57 | 0.58 |
| CT GTV + Clinics | VH.VIMP--ST | 0.56 | 0.59 | 0.58 | 0.58 |
| CT Organomics + PET Organomics | VH.VIMP--CoxPH | 0.61 | 0.56 | 0.58 | 0.58 |
| CT Organomics + PET Organomics | VH.VIMP--GLMN | 0.61 | 0.56 | 0.57 | 0.58 |
| CT Organomics + PET Organomics | VH.VIMP--RSF | 0.57 | 0.58 | 0.58 | 0.58 |
| CT Organomics + PET Organomics | VH.VIMP--GLMB | 0.61 | 0.56 | 0.58 | 0.58 |
| CT Organomics + Clinics | VH.VIMP--RSF | 0.57 | 0.56 | 0.59 | 0.58 |

| | | | | | |
|---|---|---|---|---|---|
| PET Organomics + CT Organomics + Clinics | VH.VIMP--RSF | 0.59 | 0.55 | 0.6 | 0.58 |
| CT GTV + CT Organomics | VH.VIMP--CB | 0.59 | 0.56 | 0.58 | 0.58 |
| PET GTV + PET Organomics | VH.VIMP--CB | 0.7 | 0.55 | 0.5 | 0.58 |
| PET GTV + PET Organomics | VH.VIMP--RSF | 0.6 | 0.57 | 0.58 | 0.58 |
| PET GTV + PET Organomics | VH.VIMP--ST | 0.56 | 0.56 | 0.63 | 0.58 |
| PET Organomics + CT Organomics + PET GTV + CT GTV | VH.VIMP--CB | 0.59 | 0.56 | 0.58 | 0.58 |
| PET Organomics + CT Organomics + PET GTV + CT GTV | VH.VIMP--GLMN | 0.59 | 0.55 | 0.6 | 0.58 |
| PET Organomics + CT Organomics + PET GTV + CT GTV | VH.VIMP--RSF | 0.58 | 0.56 | 0.59 | 0.58 |
| PET Organomics + CT Organomics + PET GTV + CT GTV | VH.VIMP--GLMB | 0.59 | 0.55 | 0.59 | 0.58 |
| CT GTV | UCI--CoxPH | 0.57 | 0.57 | 0.58 | 0.57 |
| CT GTV + CT Organomics + Clinics | MD--RSF | 0.59 | 0.55 | 0.57 | 0.57 |
| PET GTV + PET Organomics + Clinics | MD--ST | 0.58 | 0.55 | 0.57 | 0.57 |
| PET Organomics + CT Organomics + PET GTV + CT GTV + Clinics | MD--CoxPH | 0.59 | 0.55 | 0.59 | 0.57 |
| PET Organomics + CT Organomics + PET GTV + CT GTV + Clinics | MD--RSF | 0.58 | 0.58 | 0.56 | 0.57 |
| Clinics | MD--RSF | 0.58 | 0.56 | 0.57 | 0.57 |
| CT GTV | MD--GLMN | 0.58 | 0.56 | 0.58 | 0.57 |
| PET GTV | MD--CoxPH | 0.57 | 0.57 | 0.57 | 0.57 |
| CT GTV + Clinics | MD--GLMN | 0.56 | 0.56 | 0.57 | 0.57 |
| CT GTV + Clinics | MD--GLMB | 0.56 | 0.57 | 0.57 | 0.57 |
| PET GTV + CT GTV + Clinics | MD--GLMN | 0.59 | 0.57 | 0.57 | 0.57 |
| CT Organomics + PET Organomics | MD--GLMN | 0.56 | 0.57 | 0.58 | 0.57 |
| CT Organomics + PET Organomics | MD--GLMB | 0.57 | 0.57 | 0.57 | 0.57 |
| CT Organomics + Clinics | MD--ST | 0.6 | 0.55 | 0.57 | 0.57 |
| PET Organomics + Clinics | MD--CB | 0.63 | 0.59 | 0.5 | 0.57 |
| PET GTV + PET Organomics | MD--CB | 0.58 | 0.5 | 0.64 | 0.57 |
| CT GTV + CT Organomics + Clinics | MI--CB | 0.57 | 0.56 | 0.58 | 0.57 |
| CT GTV + CT Organomics + Clinics | MI--RSF | 0.56 | 0.56 | 0.58 | 0.57 |
| PET GTV + PET Organomics + Clinics | MI--CoxPH | 0.58 | 0.57 | 0.57 | 0.57 |
| PET GTV + PET Organomics + Clinics | MI--RSF | 0.58 | 0.56 | 0.57 | 0.57 |
| PET GTV + PET Organomics + Clinics | MI--ST | 0.57 | 0.58 | 0.56 | 0.57 |
| Clinics | MI--CoxPH | 0.56 | 0.56 | 0.6 | 0.57 |
| PET GTV + CT GTV | MI--ST | 0.58 | 0.57 | 0.56 | 0.57 |
| CT GTV + Clinics | MI--CoxPH | 0.56 | 0.57 | 0.59 | 0.57 |
| PET GTV + Clinics | MI--CoxPH | 0.56 | 0.57 | 0.58 | 0.57 |
| PET GTV + Clinics | MI--ST | 0.57 | 0.57 | 0.58 | 0.57 |
| CT Organomics + Clinics | MI--CB | 0.57 | 0.56 | 0.57 | 0.57 |
| CT Organomics + Clinics | MI--RSF | 0.59 | 0.57 | 0.57 | 0.57 |
| PET Organomics + Clinics | MI--CoxPH | 0.58 | 0.57 | 0.57 | 0.57 |

| | | | | | |
|---|---|---|---|---|---|
| PET Organomics + Clinics | MI--ST | 0.57 | 0.56 | 0.57 | 0.57 |
| PET Organomics + CT Organomics + Clinics | MI--CB | 0.57 | 0.5 | 0.63 | 0.57 |
| CT GTV + CT Organomics | MI--CoxPH | 0.58 | 0.56 | 0.58 | 0.57 |
| PET GTV + PET Organomics + Clinics | VH--CoxPH | 0.58 | 0.55 | 0.58 | 0.57 |
| PET GTV + PET Organomics + Clinics | VH--GLMN | 0.58 | 0.55 | 0.58 | 0.57 |
| PET Organomics + CT Organomics + PET GTV + CT GTV + Clinics | VH--ST | 0.57 | 0.57 | 0.56 | 0.57 |
| Clinics | VH--RSF | 0.58 | 0.55 | 0.59 | 0.57 |
| PET GTV | VH--ST | 0.58 | 0.58 | 0.56 | 0.57 |
| PET GTV + CT GTV | VH--ST | 0.56 | 0.57 | 0.57 | 0.57 |
| PET Organomics | VH--CB | 0.58 | 0.55 | 0.59 | 0.57 |
| CT Organomics + PET Organomics | VH--CoxPH | 0.58 | 0.58 | 0.57 | 0.57 |
| CT Organomics + PET Organomics | VH--ST | 0.58 | 0.56 | 0.56 | 0.57 |
| PET Organomics + CT Organomics + Clinics | VH--ST | 0.64 | 0.52 | 0.56 | 0.57 |
| CT GTV + CT Organomics | VH--ST | 0.61 | 0.56 | 0.55 | 0.57 |
| CT GTV + CT Organomics + Clinics | VH.VIMP--CoxPH | 0.57 | 0.56 | 0.58 | 0.57 |
| CT GTV + CT Organomics + Clinics | VH.VIMP--CB | 0.57 | 0.56 | 0.58 | 0.57 |
| CT GTV + CT Organomics + Clinics | VH.VIMP--GLMB | 0.57 | 0.56 | 0.59 | 0.57 |
| PET GTV + CT GTV | VH.VIMP--ST | 0.58 | 0.57 | 0.56 | 0.57 |
| PET GTV + CT GTV + Clinics | VH.VIMP--ST | 0.57 | 0.59 | 0.57 | 0.57 |
| CT Organomics + PET Organomics | VH.VIMP--ST | 0.57 | 0.57 | 0.58 | 0.57 |
| PET Organomics + Clinics | VH.VIMP--CB | 0.64 | 0.56 | 0.5 | 0.57 |
| PET Organomics + CT Organomics + Clinics | VH.VIMP--CB | 0.63 | 0.58 | 0.5 | 0.57 |
| CT GTV + CT Organomics | VH.VIMP--GLMN | 0.58 | 0.56 | 0.57 | 0.57 |
| CT GTV + CT Organomics | VH.VIMP--GLMB | 0.57 | 0.56 | 0.57 | 0.57 |
| CT GTV | UCI--CB | 0.58 | 0.5 | 0.59 | 0.56 |
| PET GTV | UCI--ST | 0.54 | 0.58 | 0.57 | 0.56 |
| CT GTV + CT Organomics + Clinics | MD--CB | 0.61 | 0.57 | 0.5 | 0.56 |
| CT GTV | MD--ST | 0.57 | 0.56 | 0.57 | 0.56 |
| CT GTV + Clinics | MD--ST | 0.58 | 0.55 | 0.56 | 0.56 |
| Clinics | MI--CB | 0.5 | 0.56 | 0.62 | 0.56 |
| CT GTV | MI--CB | 0.59 | 0.5 | 0.59 | 0.56 |
| Clinics | VH--ST | 0.56 | 0.55 | 0.56 | 0.56 |
| CT GTV + Clinics | VH--CB | 0.58 | 0.5 | 0.59 | 0.56 |
| PET GTV + CT GTV + Clinics | VH--CB | 0.59 | 0.5 | 0.57 | 0.56 |
| CT GTV + Clinics | VH.VIMP--CB | 0.58 | 0.5 | 0.6 | 0.56 |
| PET Organomics + CT Organomics + Clinics | VH.VIMP--ST | 0.56 | 0.56 | 0.57 | 0.56 |

| | | | | | |
|---|---|---|---|---|---|
| CT GTV + CT Organomics | VH.VIMP--CoxPH | 0.56 | 0.56 | 0.57 | 0.56 |
| PET GTV + Clinics | UCI--CB | 0.59 | 0.5 | 0.56 | 0.55 |
| PET Organomics + CT Organomics + PET GTV + CT GTV + Clinics | MD--CB | 0.59 | 0.56 | 0.5 | 0.55 |
| PET GTV | MD--CB | 0.58 | 0.5 | 0.56 | 0.55 |
| PET GTV + CT GTV | MD--CB | 0.58 | 0.5 | 0.58 | 0.55 |
| CT Organomics | MD--CB | 0.58 | 0.56 | 0.5 | 0.55 |
| PET Organomics + CT Organomics + Clinics | MD--CB | 0.58 | 0.57 | 0.5 | 0.55 |
| PET GTV | MI--CB | 0.58 | 0.5 | 0.56 | 0.55 |
| PET GTV + Clinics | MI--CB | 0.56 | 0.5 | 0.58 | 0.55 |
| PET GTV + CT GTV + Clinics | MI--CB | 0.58 | 0.57 | 0.5 | 0.55 |
| Clinics | VH--CB | 0.59 | 0.58 | 0.5 | 0.55 |
| Clinics | VH--GLMN | 0.59 | 0.58 | 0.5 | 0.55 |
| CT GTV | VH--CB | 0.58 | 0.5 | 0.58 | 0.55 |
| PET GTV | VH--CB | 0.5 | 0.57 | 0.57 | 0.55 |
| PET GTV + CT GTV | VH--CB | 0.58 | 0.5 | 0.57 | 0.55 |
| PET GTV + Clinics | VH--CB | 0.59 | 0.5 | 0.57 | 0.55 |
| PET Organomics + CT Organomics + PET GTV + CT GTV | VH--CB | 0.6 | 0.56 | 0.5 | 0.55 |
| PET GTV + CT GTV | VH.VIMP--CB | 0.58 | 0.5 | 0.58 | 0.55 |
| PET GTV + CT GTV + Clinics | VH.VIMP--CB | 0.59 | 0.5 | 0.57 | 0.55 |
| CT Organomics + PET Organomics | VH.VIMP--CB | 0.59 | 0.56 | 0.5 | 0.55 |
| PET GTV + PET Organomics + Clinics | MD--CB | 0.57 | 0.55 | 0.5 | 0.54 |
| Clinics | MD--GLMN | 0.57 | 0.56 | 0.5 | 0.54 |
| CT GTV + Clinics | MD--CB | 0.56 | 0.57 | 0.5 | 0.54 |
| CT GTV + Clinics | MI--CB | 0.56 | 0.56 | 0.5 | 0.54 |
| Clinics | UCI--CB | 0.58 | 0.5 | 0.5 | 0.53 |
| PET GTV + CT GTV + Clinics | MD--CB | 0.58 | 0.5 | 0.5 | 0.53 |
| PET GTV + Clinics | VH.VIMP--CB | 0.58 | 0.5 | 0.5 | 0.53 |
| Clinics | MD--CB | 0.57 | 0.5 | 0.5 | 0.52 |
| CT Organomics + PET Organomics | MD--CB | 0.5 | 0.57 | 0.5 | 0.52 |
| CT GTV + CT Organomics + Clinics | VH--CB | 0.5 | 0.57 | 0.5 | 0.52 |
